# Genetic risk effects on psychiatric disorders act in sets

**DOI:** 10.1101/2025.07.23.25332043

**Authors:** Jolien Rietkerk, Morten Dybdahl Krebs, Joel Mefford, Lianyun Huang, Kajsa-Lotta Georgii Hellberg, iPSYCH Study Consortium, Anders Børglum, Thomas Werge, Kenneth S. Kendler, Jonathan Flint, Andrew J. Schork, Andrew Dahl, Na Cai

**Affiliations:** Helmholtz Pioneer Campus, Helmholtz Zentrum München, Neuherberg, Germany; Computational Health Centre, Helmholtz Zentrum München, Neuherberg, Germany; School of Medicine and Health, Technical University of Munich, Munich, Germany; Institute for Genomics in Health, SUNY Downstate Health Sciences University, Brooklyn, NY, USA; Department of Psychiatry, University Medical Center Groningen, University of Groningen, Groningen, the Netherlands; Institute of Biological Psychiatry, Mental Health Center - Sct Hans, Copenhagen University Hospital – Mental Health Services CPH, Copenhagen, Denmark; Department of Human Genetics, David Geffen School of Medicine, University of California, Los Angeles, CA, USA; National Centre for Register-based Research, Dept. of Public Health, Aarhus University, Aarhus, Denmark; Pioneer Centre for Statistical and computational Methods for Advanced Research to Transform Biomedicine (SMARTbiomed), Aarhus University, Denmark; Department of Biomedicine, Aarhus University, Aarhus, Denmark; The Lundbeck Foundation Initiative for Integrative Psychiatric Research, iPSYCH, Denmark; Center for Genomics and Personalized Medicine, Aarhus, Denmark; Lundbeck Foundation GeoGenetics Centre, Natural History Museum of Denmark, University of Copenhagen, Copenhagen 1350, Denmark; Department of Clinical Medicine, Faculty of Health and Medical Sciences, University of Copenhagen, Copenhagen 2200, Denmark; Virginia Institute for Psychiatric and Behavioral Genetics and Department of Psychiatry, Virginia Commonwealth University, Richmond, VA, USA; Section of Genetic Medicine, University of Chicago, Chicago, IL, USA; Department of Biosystems Science and Engineering, ETH Zurich, Basel, Switzerland

## Abstract

Genetic studies of psychiatric disorders have typically assumed that all genetic effects contribute additively to disease liability. However, it is likely that psychiatric disorders have unrecognized subtypes, where synergistic sets of risk variants co-occur within certain cases more than expected under additivity. The existence of synergistic sets induces a structured form of statistical interactions called coordinated epistasis. We test for these interactions in five psychiatric disorders and find evidence for synergistic sets, and by extension, disorder subtypes. We further find that synergistic sets contributing to comorbidities are mostly disorder-specific, despite high genetic correlations between disorders, supporting current diagnostic distinctions between disorders. Finally, we find that genetic risk factors shared across disorders identify a cross-disorder subtype that is likely the result of heritable confounders, rather than disorder-specific etiology. Our results show that genetic risk effects for psychiatric disorders act in sets, implying the existence of subtypes, and re-interpret the importance of shared genetic effects in understanding disease biology and classification.

## Introduction

A central challenge in psychiatric genetics is understanding how the collective influence of numerous genetic loci, each with a small effect, culminates in disease. While thousands, perhaps even tens of thousands, of risk variants are known to exist^1^, most affected individuals carry only a small subset of these^2^. It remains unclear, and rarely investigated, whether specific risk alleles co-occur more frequently in some disease cases than others, resulting in disorder subtypes. This represents a significant gap, as psychiatric disorders have long been hypothesized to be heterogeneous^3,4^. However, prior efforts to study this heterogeneity have typically focused on defining subtypes based on phenotypic profiles rather than leveraging genetic evidence^5–9^.

To illustrate the problem we address in this paper, consider a highly polygenic disorder where the effects of risk alleles across all variants contribute additively to disease liability, with each effect size being small. Under this "additive model”^10^ we would expect all risk alleles to be equally likely to contribute to liability in any given case, and there to be no genetically heterogeneous subtypes among cases (**Figure 1A**). Conversely, when there are genetically heterogeneous subtypes, the risk alleles driving different subtypes would co-occur in cases less than expected under additivity. We term this the "antagonistic model”^10^ (**Figure 1B**). Finally, risk alleles driving the same subtype co-occur more in cases than expected under the additive model, and form a "synergistic set." We refer to this architecture as the "synergistic model”^10^ (**Figure 1C**). The existence of synergistic sets induces a structured form of statistical interactions between their polygenic effects. This can be detected using a previously established method: coordinated epistasis (CE)^10,11^, which tests for interactions between synergistic sets of unknown composition using genetic risk scores that act as their proxies (**Figure 2A**).

**Figure 1:**
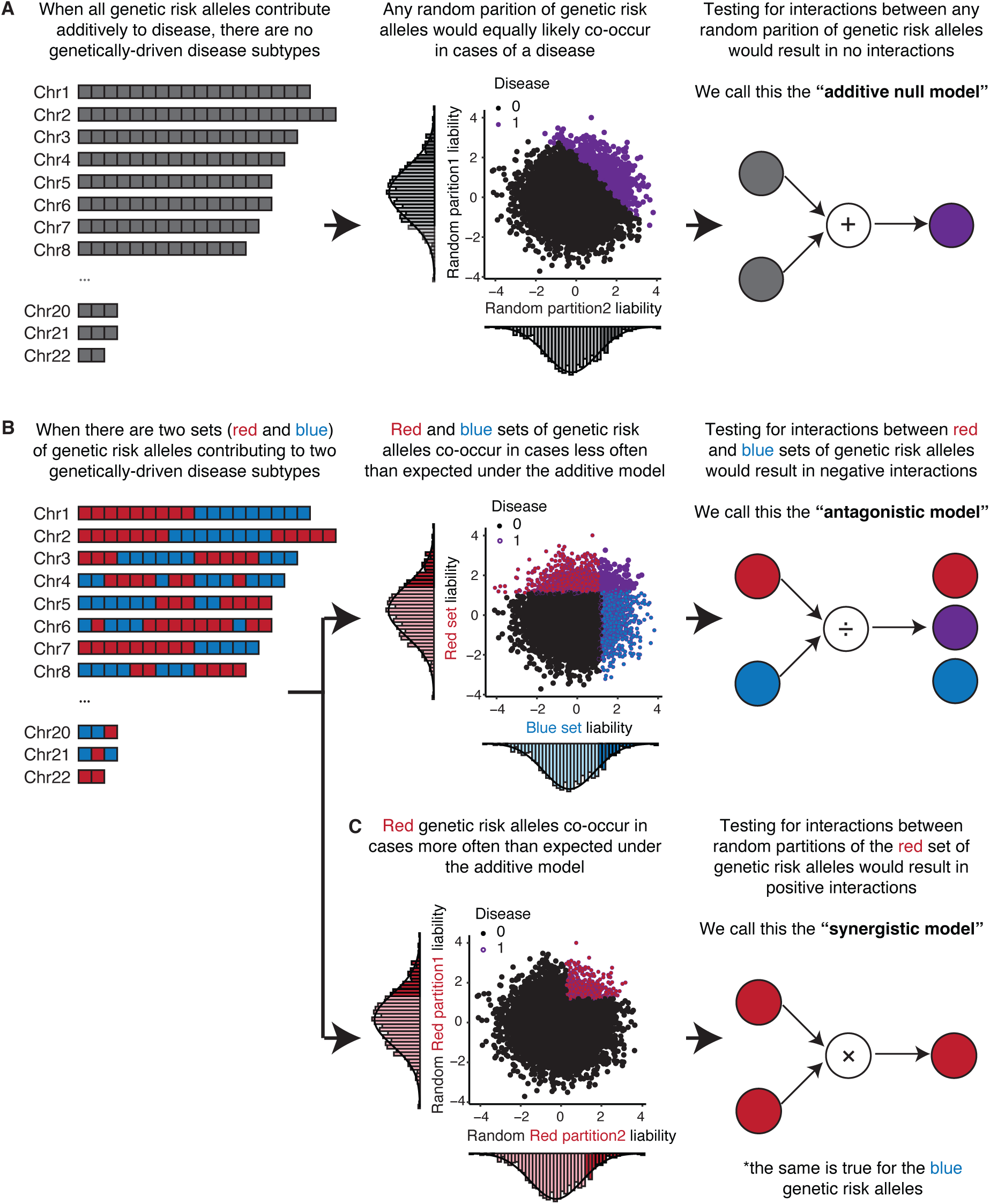
**(A)** Schematic that shows the additive “null” model: when all genetic risk effects for a disease in the genome, distributed across all chromosomes (left panel, where one square represents the risk allele at one risk variant) are additive, any random partition of the genetic risk alleles would equally likely co-occur in any case of the disease (center panel); testing for interaction in a coordinated epistasis (CE) framework between any two random partitions of risk allele would give no significant interactions (right panel). **(B)** Schematic that shows the antagonistic model: when there are different genetic risk effects for different subtypes of a disease (shown as red and blue squares respectively, left panel), they would co-occur less frequently in the same cases than expected under the additive null model (red and blue effects segregate into red and blue subtypes, center panel); testing for CE between the red and blue sets of risk effects would result in the findings of a significant negative interaction (right panel). **(C)** Schematic that shows the synergistic model: using the same example as in **(B)**, red risk effects more often occur in cases than expected under the additive model (due to the cases of the red subtype, mid-panel); testing for CE between the random partitions of the red effects would result in the findings of a significant positive interaction (right panel).

**Figure 2:**
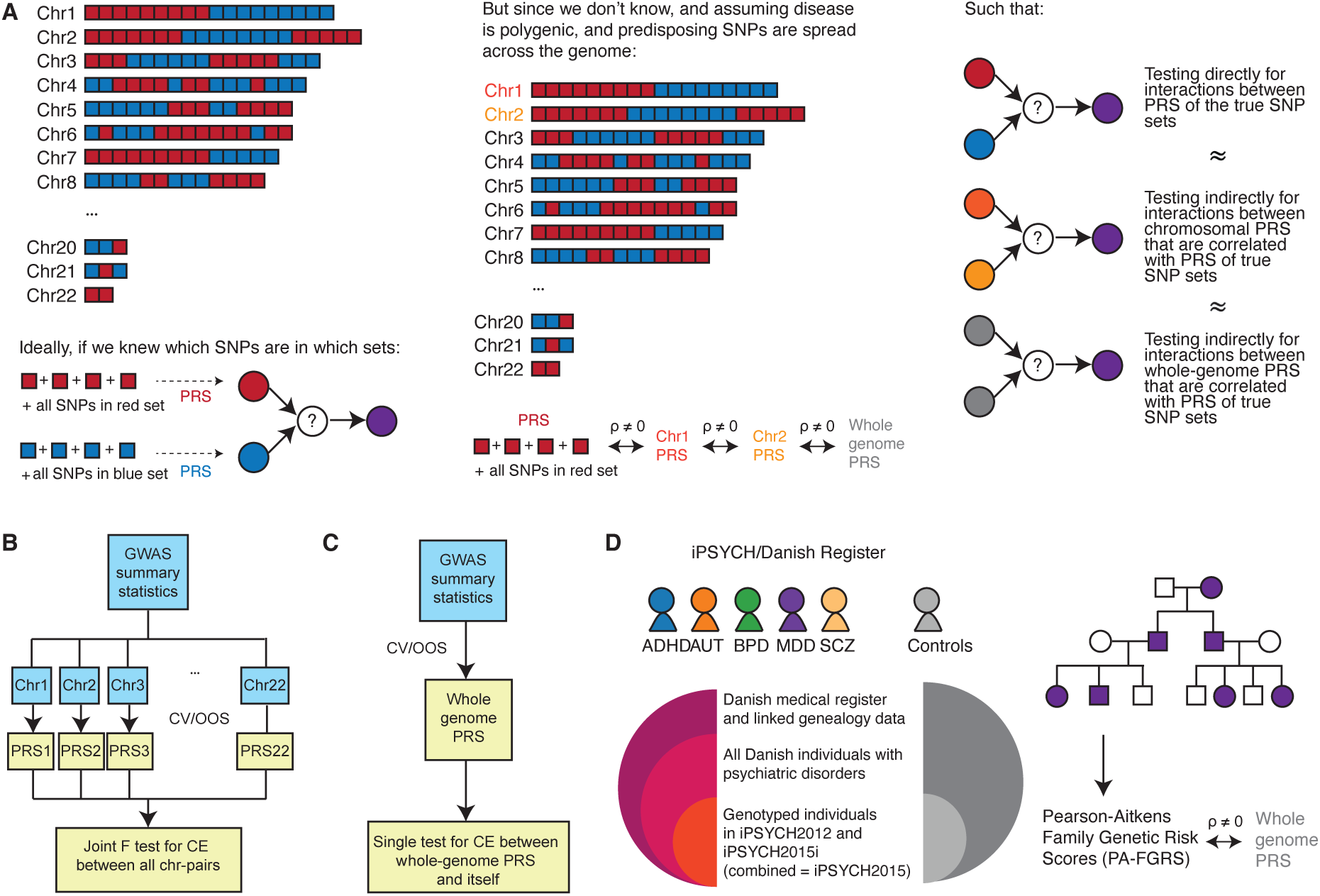
**(A)** Schematic demonstrating that ideally we would be testing for interactions, using the CE model, on risk effects that are part of the red or blue sets (left panel). However, we don’t know which risk effects belong to which set. Assuming the risk effects in the red and blue sets are distributed across all chromosomes, PRS of any chromosome or the whole genome would be correlated with PRS obtained from each of the SNP sets (middle panel), and as such CE tests performed on the chromosome partitioned PRS (pc-CE) and whole-genome PRS (wg-CE) would capturing the interactions between the red and blue sets (right panel). **(B)** Schematic for pc-CE performed using chromosomal PRS as a joint F-test across all chromosome paris; this figure shows that if pc-CE is performed in-sample, a 10-fold cross validation (CV) is performed, while if it is performed out-of-sample (OOS) then no CV is performed. **(C)** Schematic for wg-CE performed using whole-genome PRS. **(D)** Schematic of the iPSYCH cohort and Danish register data on the five psychiatric disorders; PA-FGRS obtained in iPSYCH cohorts are used to perform fgrs-CE analyses.

In this paper, we apply and extend the CE test to investigate whether multiple synergistic sets, each driving a distinct subtype, exist in five psychiatric disorders: major depressive disorder (MDD), bipolar disorder (BPD), attention deficit hyperactivity disorder (ADHD), autism spectrum disorder (AUT) and schizophrenia (SCZ). We further extend this framework to characterize the subtype architecture of comorbidity among these disorders. Finally, we examine how genetic risk shared across psychiatric disorders^12–14^ contributes to subtype architecture within and between disorders.

## Results

### Coordinated epistasis as a means to identify structure among risk effects

Coordinated epistasis (CE)^10,11^ detects the existence of multiple synergistic sets of risk variants for a disorder through testing for interactions between polygenic risk scores (PRS) computed on partitions of the genome, without the need for them to exactly reflect the compositions of the synergistic sets (**Figure 2A**). One previously proposed way to test for CE divides all risk variants by chromosome and then tests for the joint pairwise statistical epistasis between all pairs of chromosome PRS (pc-CE, **Figure 2B, Methods, Supplementary Methods**)^10^. Under the plausible assumption that synergistic sets are randomly distributed across the genome for polygenic phenotypes, this method yields an unbiased estimate of interactions between these unknown synergistic sets^10^.

The output of the CE test, the interaction effect parameter 𝛾 (gamma), quantifies the nature of the interactions induced by the existence of synergistic sets. A negative 𝛾 found between a pair of PRS indicates they contain multiple synergistic sets each driving a distinct disease subtype (**Figure 1B**). Conversely, a positive 𝛾 found between a pair of PRS indicates they contain risk variants form the same synergistic set, driving towards the same disorder subtype (**Figure 1C**). Notably, both the detection of significant positive and negative 𝛾 demonstrate the existence of synergistic sets, and by extension subtypes - the sign of 𝛾 depends on whether the pair of PRS used in the CE test contain risk variants within or across sets. If 𝛾 is zero between a pair of PRS, it indicates that all risk variants for the disorder are equally likely to contribute to liability in any given case, and hence there are no subtypes (**Figure 1A**).

In this paper, we extend the CE test in two ways. First, instead of testing CE between pairs of chromosome-based PRS of a disorder, we test the interaction between its whole-genome PRS and itself (wg-CE, **Figure 2C, Methods**). Simulations demonstrate that wg-CE improves power, as the whole-genome PRS captures more induced interaction effects between synergistic sets should they exist (**Extended Data Figure 1**). Second, to further improve power, we use Pearson- Aitkens Family Genetic Risk Score (PA-FGRS)^15^ in place of whole-genome PRS for detecting wg- CE (fgrs-CE, **Figure 2D**, **Methods**). PA-FGRS is an alternative measure of disorder liability obtained using disease status of all available relatives in a pedigree. It has been shown to explain more disorder liability than PRS^15^, especially when large pedigrees are available, as it captures non-genotyped and weak genetic effects that PRS may miss, and as such we expect it to give greater power in CE tests soo.

### CE detects non-additive genetic risk in MDD

We first investigate the existence of multiple synergistic sets of risk variants in MDD. We compute PRS for the CIDI-based MDD phenotype LifetimeMDD^16^ in UK Biobank using summary statistics from seven previously published genome-wide association studies (GWAS, **Supplementary Table S1, S2**, **Methods, Supplementary Methods)**; these include LifetimeMDD^16^ itself and six different genetic correlation (rG)^17^ weighted meta-analysis (MTAG)^18^ GWAS for LifetimeMDD^19^ in UK Biobank (**Methods**). We then perform wg-CE on LifetimeMDD using these PRS.

We find no significant pc-CE or wg-CE for LifetimeMDD using PRS derived from LifetimeMDD GWAS (through 10-fold cross validation, **Supplementary Methods**) at a 10% false discovery rate (FDR, **Methods**). However, PRS derived from MTAG.GPpsy^19^ (MTAG between LifetimeMDD and GPpsy^16^, the depression definition based on help-seeking), shows significant negative wg-CE (𝛾 = -0.020 FDR = 0.026, **Supplementary Table S3**). This could mean that MTAG.GPpsy increases power for detecting the existence of multiple synergistic sets of risk SNPs in LifetimeMDD, or that GPpsy contains synergistic sets partly distinct from those in LifetimeMDD.

To assess the robustness of these findings, we perform several simulations. First, we demonstrated that MTAG on two phenotypes simulated under the additive null model does not introduce spurious CE. Results are shown in **Extended Data Figure 2**. Next we perform MTAG adding one GWAS at a time based on each one’s absolute genetic correlation (rG, **Methods**) with LifetimeMDD. Our objective is to determine whether adding inputs with high rG with LifetimeMDD would increase power and confirm the existence of multiple synergistic sets in LifetimeMDD, implicitly asking if a high rG suggests similar synergistic set architectures. **Extended Data Figure 2** and **Supplementary Table S4** show results for adding up to 11 GWAS **(Supplementary Methods**). We find that while most family history-based depression GWAS inputs in MTAG boosts power for identifying negative 𝛾 in LifetimeMDD, other inputs do not, even those with high rG (e.g. DepAll^16^, the symptom-based definition of depression, **Supplementary Table S4**). In other words, the architectures CE tests detect are independent of rG between inputs.

Third, we assess the robustness to the choice of phenotype scale, comparing probit and linear probability models. We find that logistic regression gives fewer significant findings than other scales, verifying that it is the most conservative (**Supplementary Tables S3,S4**). Other technical concerns of performing CE within-cohort using cross-validated PRS are discussed in **Supplementary Methods** and **Extended Data Figure 3**. Overall, our results and simulations support the interpretation that MTAG increases power for detecting synergistic sets.

### CE detects non-additive genetic risk across psychiatric disorders

We then ask if synergistic sets could be identified in other psychiatric disorders. For this, we perform CE analyses on five different psychiatric disorders in two Danish registry-based cohorts: iPSYCH2012^20,21^ and iPSYCH2015i^21^ (**Methods, Supplementary Methods**): ADHD, AUT, BPD, MDD and SCZ.

For each of these five disorders, we test pc-CE and wg-CE using PRS constructed from the respective disorder’s GWAS from the Psychiatric Genomics Consortium (PGC)^22–26^ in each of the iPSYCH cohorts (**Supplementary Table S5, Methods, Supplementary Methods**). We find no significant pc-CE or wg-CE for any of the disorders (**Figure 3A**, **Supplementary Table S6**). To improve power, we again use MTAG for combining GWAS of each disease performed on one iPSYCH cohort with its respective PGC GWAS meta-analyses, and obtain PRS on the other iPSYCH cohort (**Methods**). Using these MTAG-derived PRS, we find two significant wg-CE tests at a 10% FDR: for MDD in iPSYCH2015i (𝛾 = -0.019, FDR = 0.089) and SCZ in iPSYCH2012 (𝛾 = 0.040, FDR = 0.089, **Figure 3B, Supplementary Table S6**). Notably, the 𝛾 estimate for MDD is negative, consistent with our findings using MTAG results in UK Biobank, suggesting there are multiple synergistic sets in MDD.

**Figure 3:**
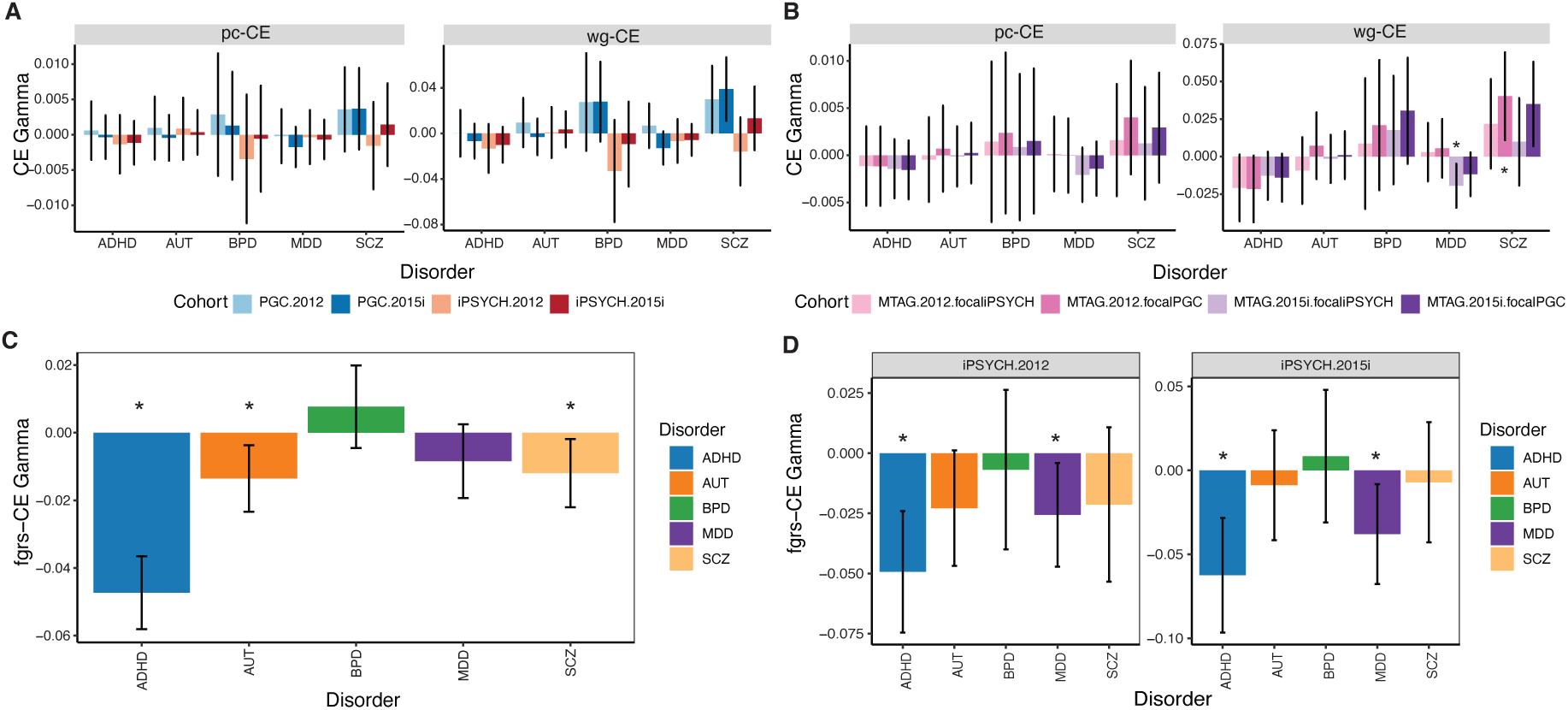
**(A)** pc-CE and wg-CE results for individual disorders, performed using PRS obtained from PGC in iPSYCH2012 (light blue) and iPSYCH2015i (dark blue), and using PRS obtained from iPSYCH2015i in iPSYCH2012 (salmon) and vice versa (red); for pc-CE, the y axis shows mean 𝛾 estimates from all chromosome pairs. **(B)** pc-CE and wg-CE results for individual disorders, performed using MTAG meta-analysis PRS between PGC and iPSYCH2015i obtained on individuals in iPSYCH2012, where the focal GWAS is iPSYCH2015i (pink) and PGC (fuschia), and using MTAG meta-analysis PRS between PGC and iPSYCH2012 obtained on individuals in iPSYCH2015i, where the focal GWAS is iPSYCH2012 (lilac) and PGC (purple). **(C)** fgrs-CE results for individual disorders in all iPSYCH individuals, performed using PA-FGRS obtained from the Danish Registry. **(D)** CE results between PRS and PA-FGRS, using PRS obtained from GWAS on iPSYCH2015i in iPSYCH2012 in the left panel, and vice versa in the right panel. For all plots, an asterisk represents FDR < 10%, error bars represent 95% confidence intervals.

To verify if our CE findings are due to cross-cohort heterogeneity, we test for cross-cohort CE between PRS derived from iPSYCH and PGC cohorts (**Methods**). We do not find evidence for this in MDD, supporting our interpretation that the consistently negative 𝛾 we identify in MDD reflects the presence of synergistic sets in MDD, rather than cross-cohort heterogeneity.

However, we observe one significant negative 𝛾 estimate between PRS derived from iPSYCH2012 and PGC GWAS for AUT^27^ in iPSYCH2015i (pc-CE mean 𝛾 = -3.77x10^-5^, FDR = 0.042, **Supplementary Table S7**). This implies that the PGC and iPSYCH2012 cohorts may have defined AUT in ways that enrich for different synergistic sets (and by extension, AUT subtypes).

### Family-based genetic risk scores improve power for CE

To further enhance our ability to detect the existence of synergistic sets in each psychiatric disorder, we obtain PA-FGRS for all individuals in the iPSYCH cohorts using their linked pedigree data (PA-FGRS^15^, **Figure 2D, Methods, Supplementary Methods, Supplementary Table S8**). Using PA-FGRS in place of whole-genome PRS in CE tests (fgrs-CE), we find that PA-FGRS significantly improves power to detect the existence of synergistic sets, as evidenced by significant negative 𝛾 estimates in fgrs-CE tests in three out of five disorders (ADHD: 𝛾 = -0.047, FDR = 1.37x10^-16^; AUT: 𝛾 = -0.014, FDR = 0.018; SCZ: 𝛾 = -0.012, FDR value = 0.030, **Figure** 3C, Supplementary Table S9).

We then investigate whether PA-FGRS captures consistent synergistic set architectures as PRS by performing CE tests between them. We perform this test in iPSYCH2012 and iPSYCH2015i separately (because the PRS for one cohort is made using GWAS derived from the other) and find significant negative 𝛾 estimates for ADHD (iPSYCH2012: 𝛾 = -0.062, FDR = 0.002; iPSYCH2015i: 𝛾 = -0.049, FDR = 0.001) and MDD (iPSYCH2012: 𝛾 = -0.038, FDR = 0.042; iPSYCH2015i: 𝛾 = -0.026, FDR = 0.049) in both cohorts (**Figure 3D**, **Supplementary Table S10**), consistent with the sign of 𝛾 identified in CE performed using PRS. These results provide further support for the existence of synergistic sets within these disorders.

### Risk effects for one disorder may be enriched in subgroups of cases of another

We next explore whether synergistic sets span disorders, given the high rGs between many pairs of disorders^28–30^, or if in fact they are distinct. To do this we introduce a new test: cross-disorder CE (cd-CE, **Figure 4A**). If synergistic sets in different disorders were distinct, we would expect to observe significant negative 𝛾 estimates between genetic scores for one disorder (D1) and another (D2) in cd-CE tests for both D1 and D2.

**Figure 4:**
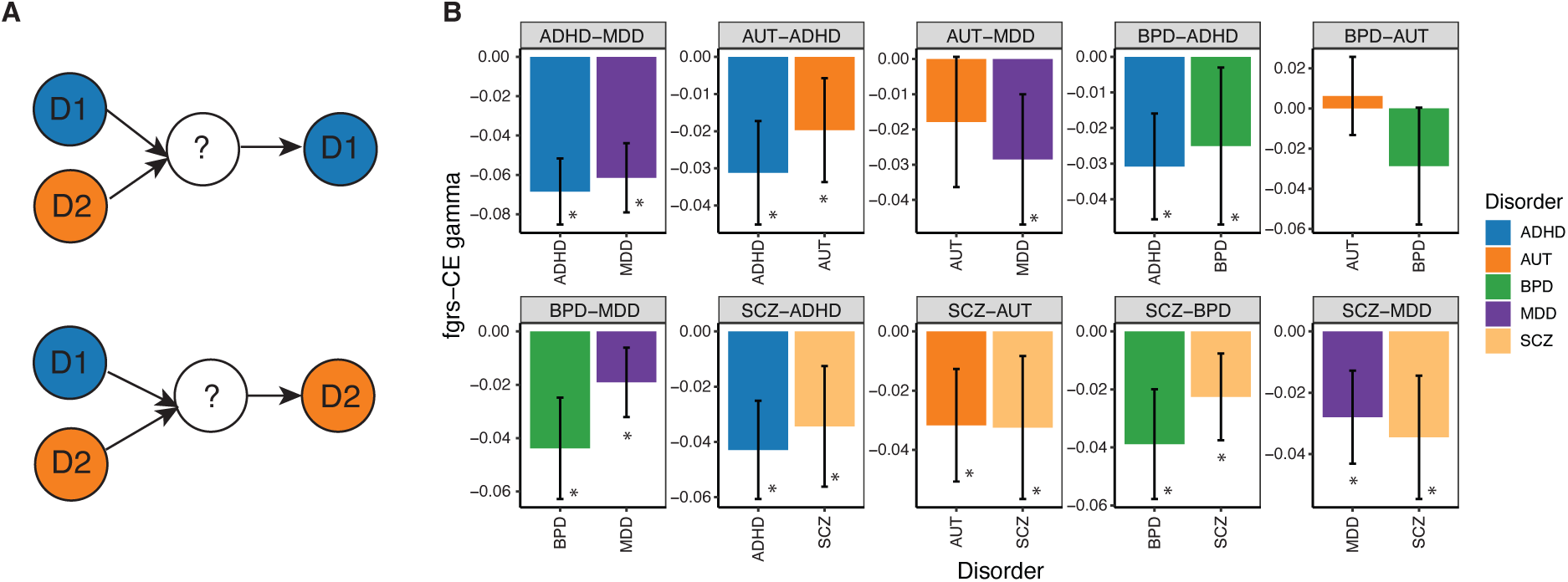
**(A)** Schematic of study design, where PA-FGRS obtained on two different disorders are tested for cd-CE for each of them. **(B)** cd-CE results using PA-FGRS for each disorder pair (panel headings) on the two constituent disorders (bars in the bar plot). For all panels, an asterisk represents FDR < 10%, error bars represent 95% confidence intervals.

We initially perform this analysis using whole-genome PRS derived from iPSYCH, PGC, and their MTAG results on pairs of disorders in each iPSYCH cohort. Consistent with the previously observed lack of power of PRS to detect CE, we find no significant cd-CE for any disorder pair (**Supplementary Table S11**). However using PA-FGRS, we find four disorder pairs with significant cd-CE (10% FDR), all of which exhibited negative 𝛾 estimates for both D1 and D2 (ADHD-MDD, ADHD-SCZ, BPD-SCZ, MDD-SCZ, **Figure 4B**). The only disorder pair where there is significant cd-CE for one of the disorders but not the other is AUT-BPD (AUT: 𝛾 = 0.0062, FDR = 0.53; BPD: 𝛾 = -0.029, FDR = 0.054, **Figure 4B, Supplementary Table S12**).

Strikingly, all significant cd-CE estimates are negative, suggesting synergistic sets do not span disorders, and are in fact distinct for each disorder (e.g. cases of ADHD and MDD are not enriched at once for both ADHD and MDD risk effects, but either one or the other). This is consistent with these disorders being etiologically distinct from each other, notwithstanding their high rG^28–30^.

### Risk effects for single disorders are enriched in subgroups of comorbid cases

We then investigate whether the liability for comorbidity between two disorders is simply an additive combination of their individual risk effects. If true, it would imply that comorbidity is a genetically distinct entity from the two individual disorders, rather than a mere co-occurrence of both.

But before that, we first perform a sanity check that CE is able to tell apart synergistic sets from two disorders when they are mixed. To do this, we define a phenotype ‘*Any’* between two disorders **(Figure 5A, Supplementary Table S13)**, where cases are individuals with both or either disorder and controls are those with neither. *Any* is therefore a “Frankenstein” phenotype, for which we would always expect negative 𝛾 in cd-CE tests between genetic risk scores from the two constituent disorders. As expected, all but one *Any* phenotype show negative 𝛾 estimates in cd-CE tests between PA-FGRS (**Figure 5B, Supplementary Table S14, S15**), confirming the cd- CE test is able to detect distinct synergistic sets for different disorders even when their cases are mixed.

**Figure 5:**
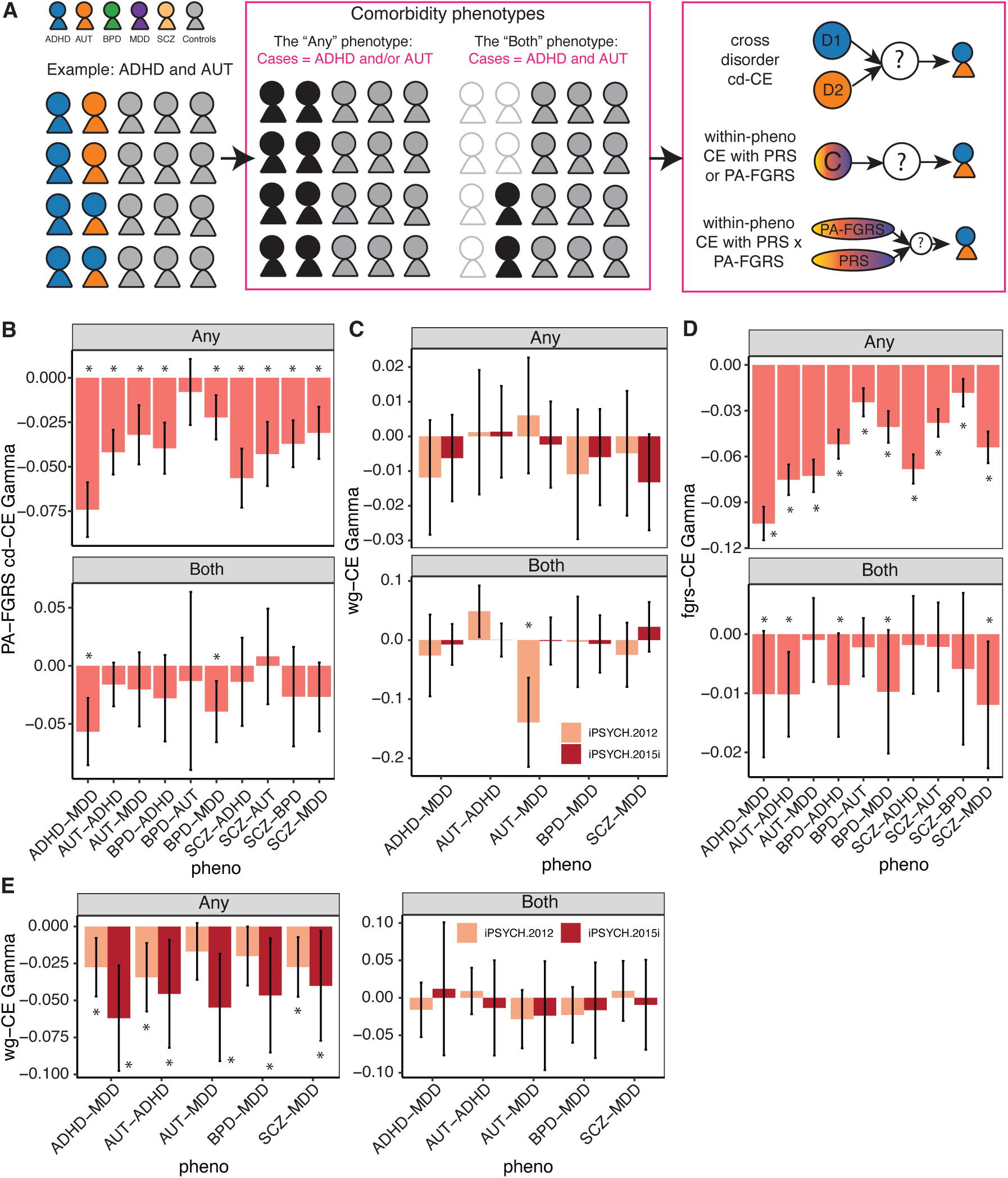
**(A)** Schematic of the two comorbidity phenotypes defined in iPSYCH, using the example of ADHD and AUT: individuals coloured in blue are cases of ADHD, individuals coloured in orange are cases of AUT, individuals with both colours are comorbid; the *Any* phenotype considers as cases (black) cases of either or both disorders; the *Both* phenotype considers as cases only those individuals who have both disorders (black), removing individuals who have either (transparent) from analysis; PRS and PA-FGRS are obtained for both comorbidity phenotypes, and then used in cd-CE and within-phenotype wg-CE or fgrs-CE tests. **(B)** fgrs-CE results cd-CE tests on the *Any* (top panel) and *Both* (bottom panel) comorbidity phenotypes, performed using PA-FGRS obtained on the whole iPSYCH2015 sample (iPSYCH2012 and iPSYCH2015i) from both constituting disorders. **(C)** wg-CE results of the *Any* (upper panel) and *Both* (lower panel) comorbidity phenotypes in five disorder pairs with cases > 250 individuals, performed using PRS from GWAS on iPSYCH2012 obtained in individuals in iPSYCH2015i (salmon) and vice versa (red). **(D)** fgrs-CE results of the *Any* (upper panel) and *Both* (lower panel) comorbidity phenotypes in five disorder pairs with cases > 250 individuals, performed using PA- FGRS obtained on the whole iPSYCH2015 sample (iPSYCH2012 and iPSYCH2015i). **(E)** Within- phenotype wg-CE results between PRS and PA-FGRS for the *Any* (left panel) and *Both* (right panel) comorbidity phenotypes in five disorder pairs with cases > 250 individuals, performed using PRS from GWAS on iPSYCH2012 obtained in individuals in iPSYCH2015i (salmon) and vice versa (red). For all panels, an asterisk represents FDR < 10%, error bars represent 95% confidence intervals.

Having ensured that, we now define a ‘*Both’* comorbidity phenotype (**Figure 5A, Supplementary Methods, Supplementary Table S13**), where individuals with both disorders are considered comorbid cases, and those with neither or only one of the two disorders are considered controls. A negative 𝛾 in cd-CE tests on the *Both* phenotype using PRS generated from the individual constituting diseases would indicate that the *Both* phenotype has subgroups of cases that are enriched in genetic effects for one of the constituting disorders. In contrast, an additive model would imply that all cases of the "*Both*" phenotype are equally likely to have their genetic effects spread evenly over both. While we find no significant cd-CE using whole-genome PRS (**Supplementary Table S14**), we observe significant cd-CE with PA-FGRS in two *Both* phenotypes, both of which have negative 𝛾 estimates (ADHD-MDD: 𝛾 = -0.06, FDR = 3.05x10^-4^; BPD-MDD: 𝛾 = -0.04, FDR =6.23x10^-4^, **Figure 5B, Supplementary Table S15**). This means comorbid cases of ADHD-MDD or BPD-MDD are specifically enriched in genetic effects for one of the two constituting disorders. This is consistent with our previous findings (**Figure 4B**) that synergistic sets do not span disorders, and support the interpretation that comorbidity is the co-occurrence of two disorders, rather than a distinct entity.

### CE tests support the current disorder delineations

Our previous analyses study the effect of disorder-specific genetic effects on comorbidity. We now turn to testing for risk effects for comorbidity using PRS and PA-FGRS (**Supplementary Methods**) constructed on *Both* and *Any* comorbidity phenotypes and performing wg-CE directly on them. Though we find no significant wg-CE (**Figure 5C, Supplementary Table S16**), our fgrs- CE results are consistent with our cd-CE findings (**Figure 5B**): all of the *Any* comorbidity phenotypes and five of the *Both* phenotypes show significant fgrs-CE, all with negative 𝛾 estimates (**Figure 5D**, **Supplementary Table S17**). These results indicate the existence of synergistic sets contributing to comorbidity, consistent with the cd-CE tests we perform in the comorbidity phenotypes (**Figure 5B**, **Supplementary Table S15**), though there are differences in statistical power.

Simulations confirm that the power difference between cd-CE and wg-CE is expected (**Extended Data Figure 4, Methods**). Specifically, cd-CE tests have higher power when the PRS from constituent disorders of a comorbidity capture synergistic sets of risk effects (**Extended Data Figure 4**). This is consistent with our results for BPD-MDD and ADHD-MDD, providing genetic support for the current clinical distinction between ADHD, BPD and MDD. Notably, our simulations show that the sign of 𝛾 estimates in cd-CE tests are consistent across a range of rGs between synergistic sets (**Extended Data Figure 4**), meaning the architecture identified by CE tests exists in addition to the high genetic correlations previously found between psychiatric disorders^28–31^.

Finally, we perform CE tests between PRS and PA-FGRS for each comorbidity phenotype to evaluate if they capture different liabilities. While significant negative 𝛾 estimates are expectedly observed for all *Any* phenotypes across the two iPSYCH cohorts, we find no significant CE between PRS and PA-FGRS for the *Both* phenotypes (**Figure 5E**, **Supplementary Table S18**). As such, we find no evidence that PRS and PA-FGRS capture different contributions to comorbidity liabilities.

### Shared genetics between disorders likely index common confounders

Finally, we utilize CE to examine how shared effects between psychiatric disorders contribute to the genetic architecture of individual psychiatric disorders and their comorbidities. Using genomic structural equation modelling (gSEM^29,32–34^), we obtain estimates of a genetic factor common to five psychiatric disorders (**Methods**, **Figure 6A**). We build a PRS for this common factor in both iPSYCH2012 and iPSYCH2015i (**Methods**), and then perform a wg-CE test using this common factor PRS in the five individual disorders in the two iPSYCH cohorts.

**Figure 6:**
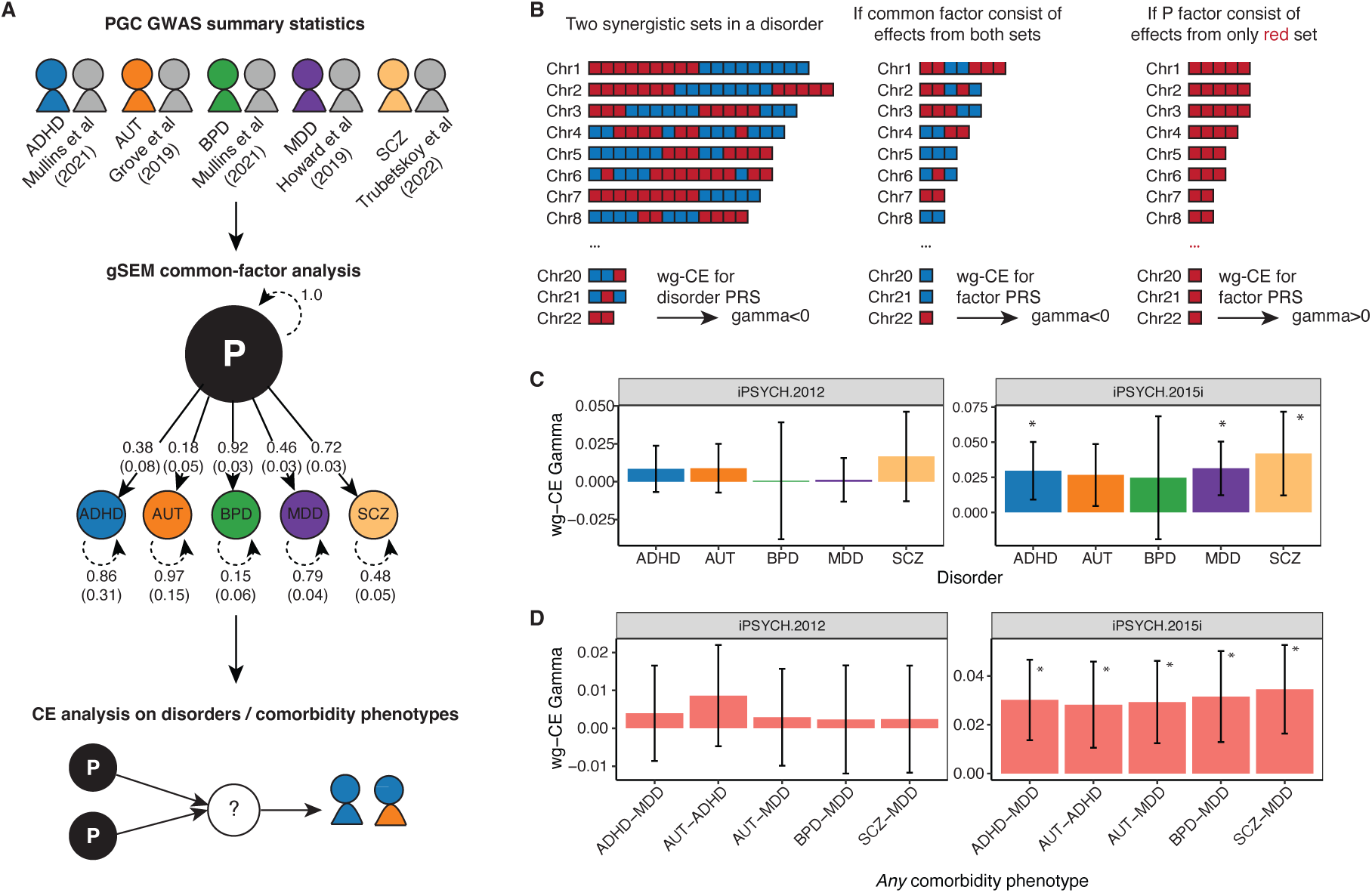
**(A)** gSEM common factor analysis to obtain a common P factor analysis on PGC GWAS on five disorders, where the P factor loads differently on each input disorder; GWAS on the P factor is then used to obtain PRS in iPSYCH2012 and iPSYCH2015i cohorts to perform pc-CE and wg-CE analyses in the respective cohorts on both individual disorder and comorbidity phenotypes. **(B)** wg-CE results obtained using P factor GWAS on each disorder in iPSYCH2012 (left panel) and iPSYCH2015i (right panel). **(C)** wg-CE results obtained using MTAG focal GWAS for each disorder on the whole iPSYCH2015 sample (iPSYCH2012 and iPSYCH2015i), on each of the five disorder-pairs with cases > 250 individuals for the *Any* (left panel) and the *Both* comorbidity phenotypes (right panel). **(D)** wg-CE results obtained using P factor GWAS on the whole iPSYCH2015 sample (iPSYCH2012 and iPSYCH2015i), on each of the five disorder-pairs with cases > 250 individuals for the *Any* (left panel) and the *Both* comorbidity phenotypes (right panel). For all panels, an asterisk represents FDR < 10%, error bars represent 95% confidence intervals.

Our previous findings show that each psychiatric disorder contains multiple synergistic sets of genetic risk effects. If the shared genetic effects between disorders, captured by this common factor, are components of these synergistic sets, then wg-CE test on the common factor PRS should yield significant negative 𝛾 estimates (**Figure 6B**, **Extended Data Figure 4**). Conversely, if the common genetic factor does not capture components of different synergistic sets, but one in particular, we would expect to see significant positive 𝛾 estimates in wg-CE tests (**Figure 6B, Extended Data Figure 4**). We find significant positive 𝛾 estimates in wg-CE tests on the common factor PRS in three out of five disorders in iPSYCH2015i (**Figure 6C**, **Supplementary Table S19**), consistent with the latter scenario. Using the same logic, we perform the wg-CE test for the common factor PRS in comorbidity phenotypes (**Supplementary Table S13**). We find significant wg-CE with positive 𝛾 estimates on all of the *Any* phenotypes in iPSYCH2015i (**Figure 6D, Supplementary Table S21**). Since the *Any* phenotypes are simply “Frankenstein” mixtures of individual disorders, the common factor behaves similarly here as it does in wg-CE analyses on the individual disorders. Both analyses suggest the common factor PRS captures effects from a single synergistic set rather than multiple, and drives a cross disorder subtype.

In supplementary materials we show results for testing for cd-CE between the common factor PRS and individual disorder PRS (**Supplementary Table S20**) and the common factor PRS for the *Both* phenotypes (**Supplementary Table S21**). Neither test is significant, likely due to lack of power.

## Discussion

Our findings reveal that the genetic risk effects for psychiatric disorders are not purely additive. Instead, we demonstrate the existence of multiple synergistic sets of risk effects that co-occur in certain disorder cases more frequently than predicted by an additive model, suggesting there are hidden subtypes in each disorder. Our current method, however, does not identify the specific variant composition of these synergistic sets. Nonetheless, our results inform their properties: in addition to showing they exist within disorders and drive hidden subtypes, we show that they are disorder-specific, and do not merge to form an additive comorbidity liability, supporting current nosological distinctions. These conclusions directly oppose the long-standing conjectures, based on high genetic correlations between psychiatric disorders and an assumption of additivity across all risk effects, that high comorbidities are expected between psychiatric disorders and current clinical boundaries do not reflect their underlying pathogenic processes^28^.

One implication of these synergistic sets, which represent coherent etiological entities, is that they may be used to define disease subtypes based on genetic etiology. This will be most fruitful if synergistic sets can be mapped to distinct and interpretable mediators such as epigenetic, transcriptomic, or proteomic signatures^35^ or clinical profiles such as symptoms or treatment response. However, determining the precise relationship between synergistic sets and clinically detectable subtypes requires knowledge of their genetic composition. Future work could involve partitioning whole-genome effects into pre-defined sets and testing if they fit the properties of a synergistic set. Although the search space for these sets is combinatorially vast, progress may be made by partitioning the genome based on pleiotropic effects or functional genomic annotations^36–41^.

A second implication of our results is that genetic correlation overstates the etiological relationship between psychiatric disease. For a decade it has been postulated that profiling and factorizing genetic correlation between disorders will identify their shared and unique etiologies, thereby improving psychiatric nosology^12,28,29,33^. However, our CE analyses demonstrate that this approach is insufficient in two key ways.

First, our discovery that the liability to comorbidity between two disorders is not necessarily an additive combination demonstrates that comorbidity does not simply depend on genetic correlation^28,33,42^. For example, our CE results show that comorbid cases of two disorders (e.g., MDD and ADHD) do not, in fact, have their comorbidity liability evenly distributed across genetic effects leading to both disorders. This observation may explain the inconsistency between observed rates of comorbid psychiatric disease within families^43–45^ and genetic correlations estimated from SNP data^12,29,46^.

Second, our findings indicate that genetic correlation is insufficient for informing nosology^28^, as high genetic correlations between disorders does not imply that they share a synergistic set that corresponds to a common etiological entity. Rather, the CE test provides a metric for determining whether two putative disorders (or subtypes of a single disorder) should be split or merged based on the overlap of their synergistic sets while accounting for genetic correlation^28–31^. For instance, the broad antagonistic CE observed across disorders within iPSYCH supports their current nosological distinction. A potential path forward is to use CE to evaluate current DSM stipulations for inclusion and exclusion of comorbid cases (e.g., MDD cases must not have BPD). Additive or synergistic cd-CE between disorders would suggest they share a synergistic set of genetic effects, thereby supporting inclusion. Conversely, antagonistic cd-CE would suggest they are driven by different synergistic sets, supporting exclusion. Similarly, CE can inform whether existing disorders should be further divided into etiologically distinct subtypes.

A third implication arising from our work concerns the interpretation of a common genetic factor underlying multiple psychiatric diseases. Several studies have interpreted these common factors as important etiological axes^13,29,32,47,48^. Our results present an alternative, plausible interpretation: the common genetic factor across five psychiatric disorders indexes a synergistic set that most likely originates from a common heritable factor external to the etiology of each disorder, such as a confounder in diagnostic protocols. For example, if diagnoses for each disorder are derived from self-administered questionnaires, they would all be confounded by heritable features related to how individuals complete questionnaires^49–51^—which are unrelated to the etiology of the disorder.

Our paper extends the CE framework^10,11^ from quantitative phenotypes to binary phenotypes like psychiatric disorders. Mathematically, this simply replaces linear regression with generalized linear models. Conceptually, however, CE for binary traits is more similar to methods for genetic subtyping. For example, our cd-CE test is similar to BUHMBOX^52^, which tests for a misdiagnosis-based subtype by asking if risk effects for another disease cluster within cases. More broadly, the CE test is a polygenic extension of "mutual exclusivity" and "multi-hit" models used to classify cancer subtypes^53^, which correspond to our antagonistic and synergistic models, respectively. The novelty of our approach lies in its agnosticism: it characterizes subtypes at the level of induced genetic interactions rather than relying on an explicit phenotypic model. While this nonparametric approach comes at a cost in power, it offers greater flexibility and robustness and, critically, enables analyses in scenarios where the underlying subtype architecture is unknown.

Several considerations should be borne in mind when interpreting our results. First, most of our significant findings originate from CE performed on PA-FGRS, which add substantial power over PRS but may also capture non-genetic familial and environmental effects, as well as assortative mating^54,55^. Nonetheless, such non-genetic contributions are likely to be small^15^ (**Supplementary Methods**) and the CE test has been shown to be robust to these effects^10^. Second, in theory, interaction effects can depend on the phenotype measurement scale, which manifests in binary phenotypes as the choice of link function in a generalized linear model. Despite this, scale is unlikely to affect our conclusions, as we examined robustness across two additional scales (linear probability and probit). Third, our analyses are limited to European genetic ancestries and to phenotyping practices in the UK Biobank and iPSYCH. It is probable that our results and conclusions, particularly concerning confounders and misdiagnosis, are partly specific to these cohorts, although our use of PRS derived from PGC GWAS mitigates this concern. With the increasing availability and use of Electronic Health Records (EHRs) and medical registries from around the world in genetic research^56–61^, we anticipate being able to utilize more diverse datasets to derive comorbidity phenotypes, thereby allowing for broader investigations into how individual disorder etiologies relate to one another.

Overall, our work has shown that genetic effects on psychiatric disorders act in synergistic sets using an approach which offers a metric for identifying the existence of distinct etiological entities, with implications for improving psychiatric diagnostics and nosology. Though our work is focused on psychiatric disorders, this approach is widely applicable to all complex traits and diseases, especially where clinical diagnostic boundaries are unclear and subtype architectures are unknown.

## Methods

### GWAS in UK Biobank and iPSYCH cohorts

For UK Biobank, GWAS was performed using imputed genotype data at 5,776,313 SNPs (MAF >= 0.05, INFO score >= 0.9) in PLINK2^62^. We used 20 PCs computed with flashPCA^63^ on independent SNPs from 337,198 White-British individuals in UK Biobank and genotyping arrays as covariates (**Supplementary Methods**), using a logistic regression model for binary phenotypes. For iPSYCH, we performed GWAS using logistic regression in PLINK2^62^ on MDD defined by at least one specialty psychiatric care contact registered in the Danish Psychiatric Central Research Register (PCR)^64^ or the Danish National Patient Register (DNPR)^65^ for ICD10 code of F33, in two independent iPSYCH cohorts iPSYCH2012^20^ and iPSYCH2015i^21^: with 42,250 and 23,351 unrelated individuals with European genetic ancestry respectively. We used 25 genomic PCs from individuals in iPSYCH2012 and iPSYCH2015i as covariates to control for population structure in each of the cohorts. Details of the iPSYCH cohorts can be found in **Supplementary Methods**.

### Genetic correlation (rG) and genomic structural equation modelling (gSEM)

To obtain rG between pairs of phenotypes in UK Biobank, we use LD score regression implemented in LDSC v1.0.1^66^, using in-sample LD scores estimated from 10,000 random White British UK Biobank^67^ individuals at SNPs with MAF > 0.05 as reference. We use the genetic covariance matrix obtained from LDSC as input in Genomic SEM^32^ (gSEM) to obtain a common factor GWAS using the commonfactorGWAS() function in the GenomicSEM R package.

### Meta-analyses

We perform meta-analysis between individual disorder GWAS performed in either iPSYCH cohorts and previously published GWAS of the same disorders conducted by the respective PGC Working Groups (**Supplementary Table S5**) using MTAG^18^. MTAG summary statistics for each focal disorder are then used for CE analyses. In particular, to investigate the influence of increasing complexity of meta-analyses on the CE outcome, as well as the influence of individual GWAS inputs into a meta-analysis on CE outcomes, we use a novel approach were MTAG is performed using increasing number of input GWAS summary statistics (incrementalMTAG, **Extended Data Figure 2**), ordered by their absolute rG with the focal phenotype of interest.

### Polygenic risk scores (PRS)

Out-of-sample PRS are obtained for individuals in each iPSYCH cohort using external GWAS or GWAS meta-analysis summary statistics as shown in **Supplementary Tables S1, S2** and **S5**. We use 25 cohort-specific genomic PCs as covariates for all PRS obtained in the iPSYCH cohorts (**Supplementary Methods**). Within-sample PRS obtained for individuals in UK Biobank using GWAS performed in UK Biobank are obtained in a 10-fold cross validation (CV) manner, where GWAS is performed in 90% of individuals and PRS obtained in the remaining 10% in each fold. To correct for artefacts (discussed in **Supplementary Methods**) we then perform a Mundlak correction^68,69^ on the obtained PRS. We use 20 genomic PCs and the genotyping array as covariates for all PRS obtained in UK Biobank (**Supplementary Methods**). For both our-of-sample and within-sample PRS, we compute the PRS in target individuals using PRSice-2^70^ with options --bar-levels 1 and --fast-score on clumped SNPs obtained from each reference GWAS using --clump-kb 250kb --clump-p 1.000000 --clump-r2 0.1. PRS are normalized to mean 0 and sd 1 prior to application into the CE framework using the scale() function in R.

### Pearson-Aitkens Family Genetic Risk Scores (PA-FGRS)

For individual disorder CE tests and cross-disorder CE tests that use individual disorder PA-FGRS, scores are obtained from a previous study^15^. For comorbidity phenotypes, PA-FGRS scores are obtained using a kinship matrix generated from genealogy data in the Danish Register using the PA-FGRS R package assuming a heritability of 0.4 for all comorbidity phenotypes, which includes phenotype information from affected family members (**Supplementary Methods**). PA-FGRS are directly used within the CE framework after a normalization to mean 0 and sd 1 using the scale() function in R.

### Coordinated Epistasis (CE) tests

We perform three types of tests for CE in this manuscript: per-chromosome CE (pc-CE), whole-genome CE (wg-CE) and CE between PA-FGRS (fgrs-CE). For both pc-CE and wg-CE, PRS are obtained out-of-sample for iPSYCH cohorts, and in-sample using 10-fold CV for UK Biobank with Mundlak correction (**Supplementary Methods**). In pc-CE we model:

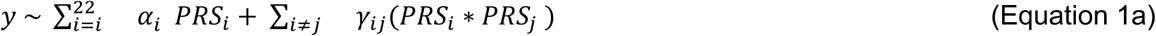

where *y* is a phenotype, 𝑃𝑅𝑆_𝑖_ is the PRS built from SNPs on chromosome *i*, and * indicates element-wise multiplication We test for CE using an F-test of the null hypothesis that all PRS interactions are zero (𝛾_𝑖j_ = 0 for all *i* and *j*). In the main text, we focus on the mean 𝛾_𝑖j_, the genome-wide CE. In wg-CE and fgrs-CE, we model:

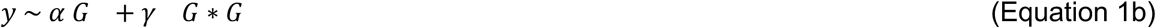

and perform a t-test for 𝛾 ≠ 0, where 𝐺 is either whole-genome PRS or PA-FGRS. We also adapt this approach to test for interactions between two different scores, such as the PRS for two distinct disorders or the PRS for the P factor, which we write as:

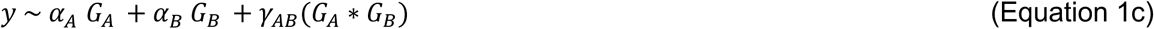

We call this cross-disorder CE (cd-CE). Notably, the phenotype 𝑦 here can be phenotypes 𝐴, 𝐵, or their comorbidity phenotype *Any* or *Both*. We primarily use logistic regression to fit these regression equations, though we also perform robustness tests by instead using linear or probit regressions.

### Simulations

First, we perform simulations to ask how pc-CE and wg-CE compare in terms of power and ability to capture ground truth 𝛾, as shown in **Extended Data Figure 1**. To do this we model the phenotype 𝑦 as a quantitative trait under our previously published model of interaction between latent liabilities^10,11^:

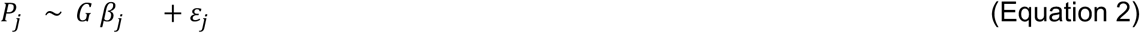

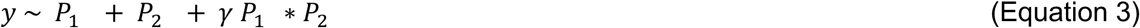

where 𝑃_j_ is a latent liability, which can be understood as genetic effects leading towards a subtype *j* of a disease liability 𝑦, 𝐺 is a genotype matrix (assumed i.i.d. standard normal for simplicity) and 𝜀_j_ is i.i.d Gaussian noise with variance chosen such that the total variance of 𝑦 is 1. We use N=20,000 individuals, M=100 causal SNPs, additive heritability 0.2, and we vary 𝛾 (gamma). We perform GWAS on simulated 𝑦 in 10000 training individuals to obtain effect sizes β (beta) at all M SNPs, construct PRS in the 10000 held-out test individuals using all 100 SNPs and random 50% splits of the 100 SNPs^10^ (pseudo-chromosomes). We then test for wg-CE using PRS built from all 100 SNPs and pc-CE using PRS built from the pseudo-chromosomes, and obtain estimates of 𝛾 (gamma-hat).

Second, we perform simulations under an additive multi-trait model to evaluate whether MTAG induces false positive CE, as shown in **Extended Data Figure 2a**. We simulate:

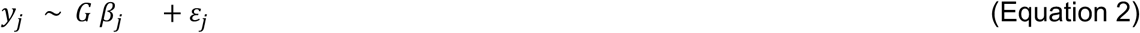

where *j* indexes phenotypes. We simulate *j*=4 phenotypes with 70% genetic and 70% nongenetic correlation across them, and perform MTAG on the effect sizes obtained from GWAS on 𝑦_1_ to 𝑦_4_ . We then arbitrarily divide the 100 causal SNPs into 20 partitions, and obtain partition-PRS for both the ordinary GWAS for 𝑦_1_ as well as the MTAG GWAS treating 𝑦_1_ as the focal trait. We perform 6 replicate simulations; for each simulation replicate, we perform pairwise interaction tests (equivalent to pc-CE) on the partition-PRS, and find no significant interaction effects (Extended Data Figure 2a). Our results remain calibrated for a range of tested values for genetic and non-genetic correlations between phenotypes, including scenarios where genetic and nongenetic correlation have opposite signs.

Third, we perform a simulation using the same model for 𝑃_j_ as Equation 2, but now the pathways are latent liabilities that define disease by one of three liability threshold models:

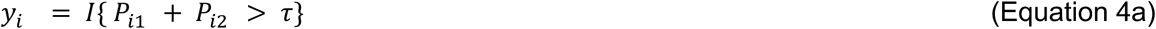

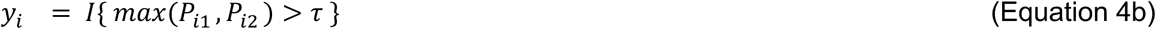

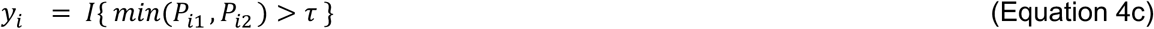

where *I{.}* is a binary indicator function and 𝜏 is defined such that 𝑦 has prevalence 30%. The first model is additive: disease depends only on the sum of the two liabilities. The second is a subtype model: many cases only have high liability on one pathway. The third is a synergistic model: all cases must have high liability on both pathways. We particularly focus on the second model in this paper.

We first evaluate whether as 𝑃_1_ ∗ 𝑃_2_ cd-CE test is better powered than wg-CE test 𝑦_𝑖_ under the antagonistic model, where liabilities 𝑃_1_ and 𝑃_2_ can be understood as those driving two subtypes of a disorder 𝑦_𝑖_, as shown in **Extended Data Figure 4**. Note that in practice the latent subtypes may not be well-identified.

We then ask what wg-CE on individual latent liabilities would show in the antagonistic subtype model. To do this we perform the above 𝑃_1_ ∗ 𝑃_2_ cd-CE test as well as wg-CE tests for each latent liability (𝑃_1_ ∗ 𝑃_1_ and 𝑃_2_ ∗ 𝑃_2_) for the antagonistic subtype model, as shown in **Extended Data Figure 4**. Note here we vary the genetic correlation between 𝑃_1_ and 𝑃_2_, to demonstrate that the signs of interactions CE identify are consistent regardless of the genetic correlations between them, though power decreases as their genetic correlation increases.

## Data Availability

UK Biobank genotype and phenotype data used in this study are from the full release (imputation version 2) of the UK Biobank Resource obtained under application no. 28709 and 163937. We used publicly available summary statistics from PGC29 and 23andMe from the Psychiatric Genomics Consortium (https://www.med.unc.edu/pgc/results-and-downloads), with references in Supplementary Table S5. Individual-level Danish data is not publicly available due to institutional restrictions on data sharing and privacy concerns. Summary statistics of the GWAS on the gSEM common factor are available at: https://doi.org/10.6084/m9.figshare.28652153.

## Extended Data Figures

**Extended Data Figure 1:**
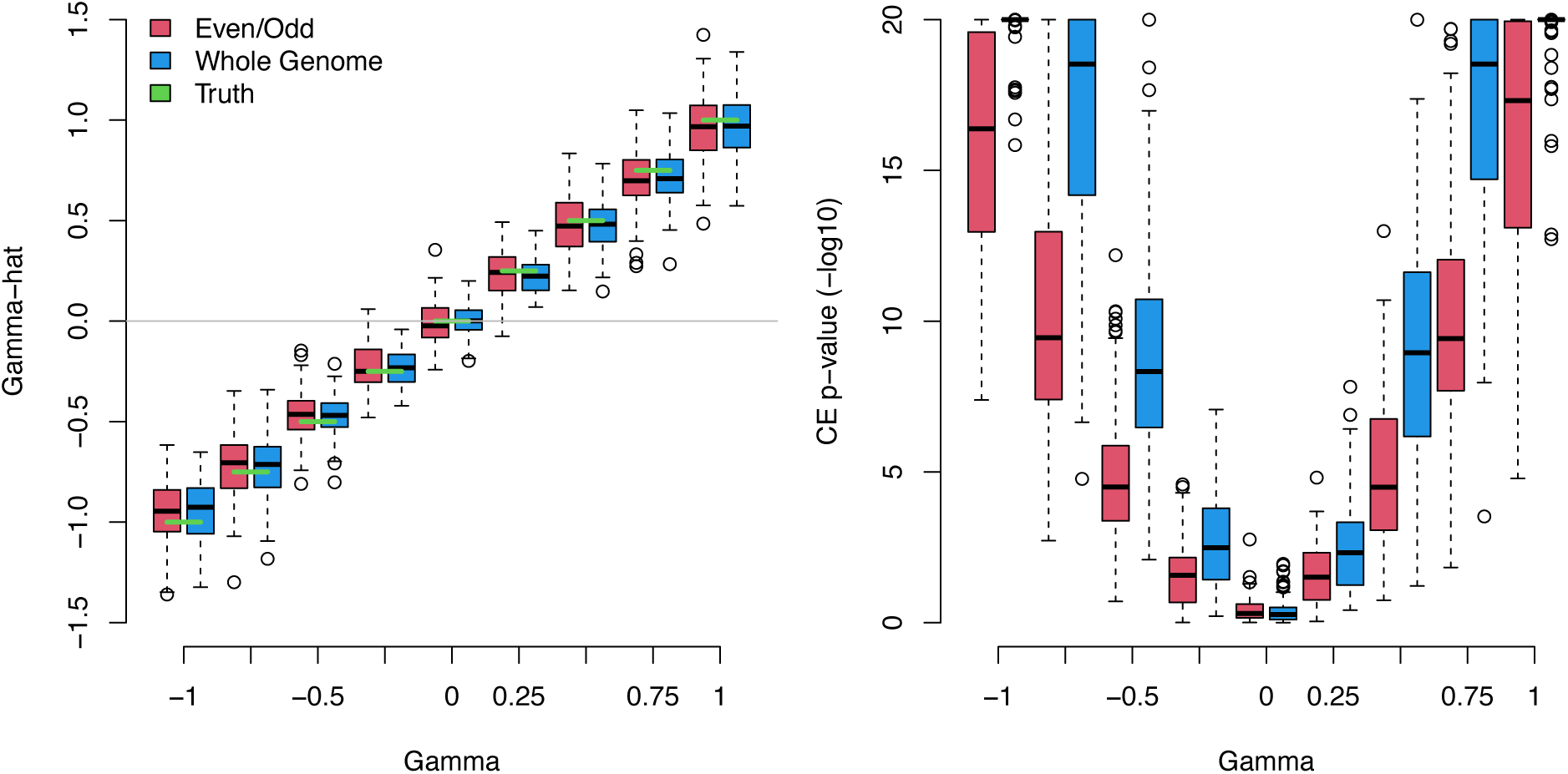
(Left panel) Simulated 𝛾 (Gamma, x axis) plotted against observed 𝛾 (Gamma-hat, y axis) in CE tests across 100 simulations of 2-pathway interactions across different gamma values between -1 and 1; green lines show simulated 𝛾, red boxplots show observed 𝛾 values in half-genome (even-odd, EO) CE tests performed on the 100 simulations, blue boxplots show observed 𝛾 values in wg-CE tests performed on the 100 simulations; both EO and wg-CE tests are able to identify simulated 𝛾. (Right panel) simulated 𝛾 (Gamma, x axis) plotted against CE test -log10(P values); red boxplots show the EO-CE test -log10(P values), blue boxplots show the EO-CE test -log10(P values); blue boxplots are always higher than red boxplots, showing wg-CE has higher power than EO-CE or, by extension, partitioned genome CE such as pc-CE. For all boxes, upper and lower hinges represent the first and third quartiles of the values, the whiskers extend to the largest value smaller than 1.5 times the interquartile range, and data points beyond the interquartile range are plotted as empty dots beyond the whiskers.

**Extended Data Figure 2:**
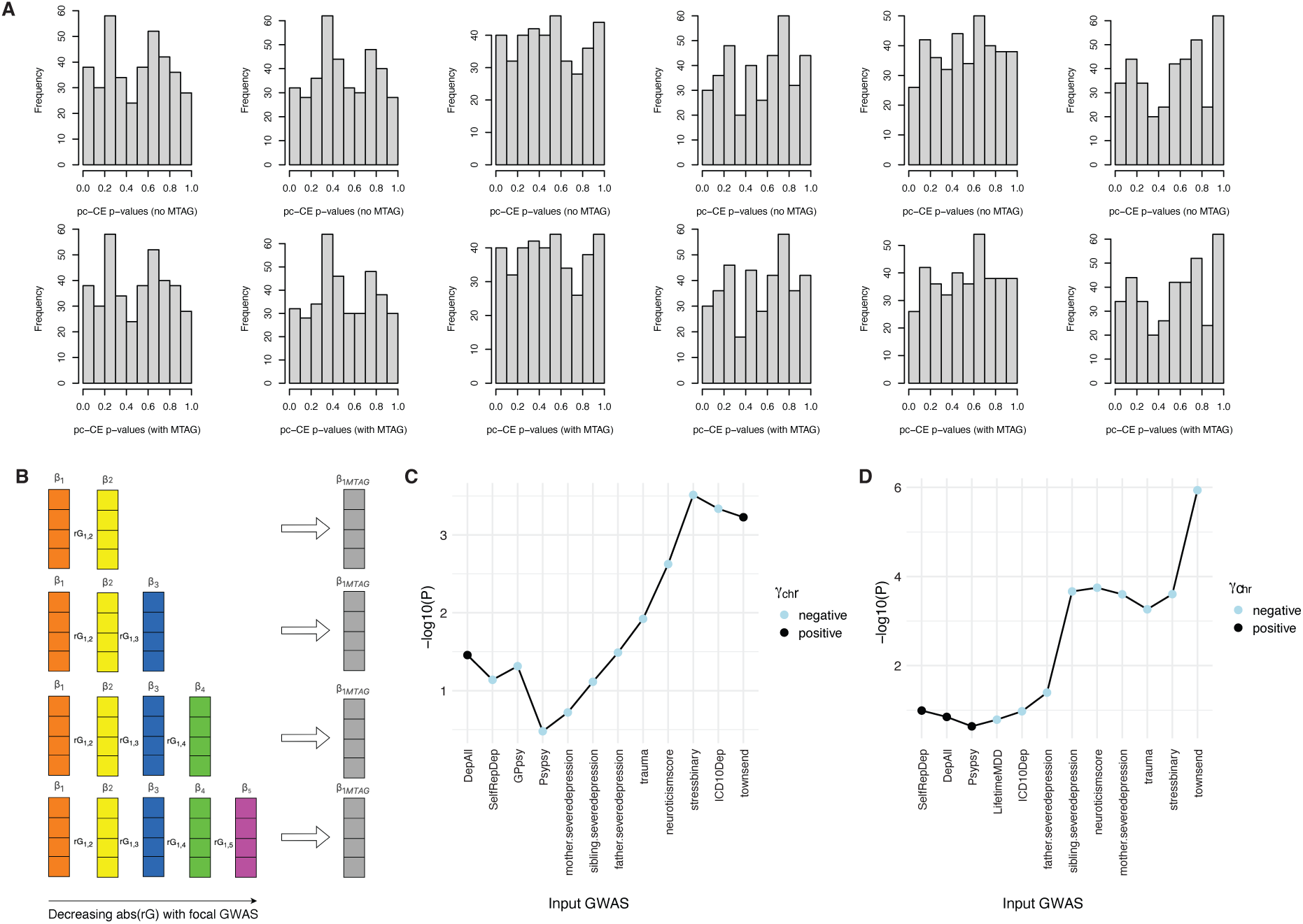
**(A)** Six replicate simulations show calibrated null P values for testing cross-chromosome CE without (top) and with (bottom) MTAG under an additive model, demonstrating MTAG does not introduce spurious CE. **(B)** Schematic of incrementalMTAG, where MTAG is performed using increasing numbers of input GWAS ranked based on their absolute rG with the focal phenotype. **(C)** pc-CE -log10(P values) using PRS from steps of incrementalMTAG on all input GWAS of MTAG.All, where the focal phenotype is LifetimeMDD; blue dots represent pc-CE results with negative mean γ, black dots represent pc-CE results with positive mean γ. **(D)** pc-CE -log10(P values) using PRS from steps of incrementalMTAG on all input GWAS of MTAG.All, where the focal phenotype is GPpsy; blue dots represent pc-CE results with negative mean 𝛾, black dots represent pc-CE results with positive mean 𝛾.

**Extended Data Figure 3:**
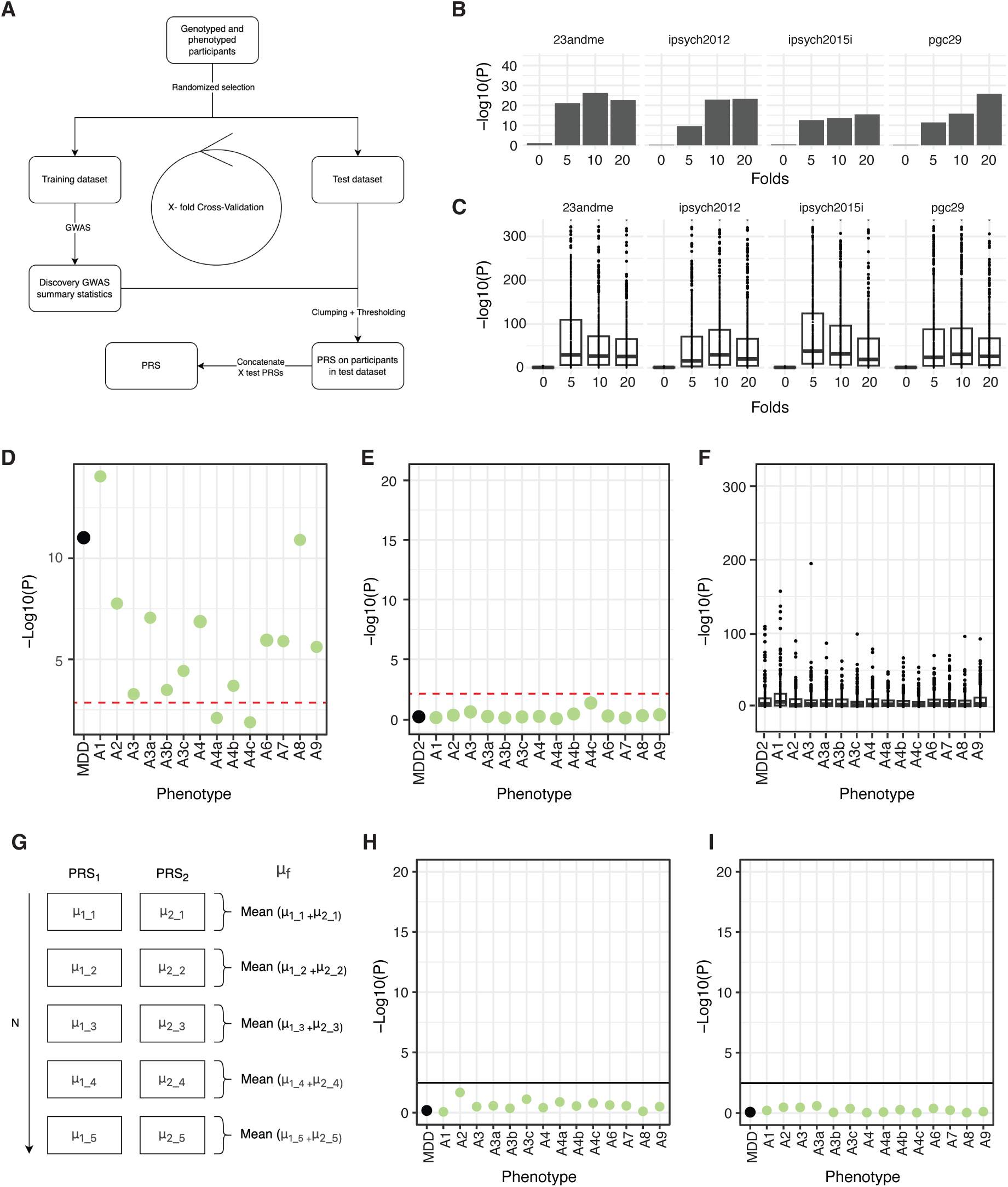
**(A)** Schematic of how within-cohort PRS is obtained using 10-fold cross validation (CV). **(B)** The effect of number of folds of external GWAS trained PRS (on UK Biobank individuals) on pc-CE -log10(P values), where the P value threshold for PRS training is not fixed; the greater the number of folds, the greater the batch effect created by the floating P-value threshold for PRS training, the greater the inflation in pc-CE -log10(P values). **(C)** The effect of number of folds of external GWAS trained PRS (on UK Biobank individuals) on Pearson correlation -log10(P value) between PRS on all pairs of chromosomes; the greater the number of folds, the greater the artefactual correlation between PRS on pairs of chromosomes. **(D)** pc-CE - log10(P values) on LifetimeMDD and CIDI-SF derived symptoms of MDD in UK Biobank, when PRS is obtained using 10-fold CV, where the P-value threshold for PRS training is not fixed; this shows a great inflation of pc-CE -log10(P values). **(E)** pc-CE -log10(P values) on LifetimeMDD and CIDI-SF derived symptoms of MDD in UK Biobank, when PRS is obtained using 10-fold CV, where the P-value threshold for PRS training is fixed at 1; this shows a great reduction in CE - log10(P value) inflation. **(F)** Pearson correlation -log10(P value) between chromosomal PRS on LifetimeMDD and CIDI-SF derived symptoms of MDD in the UK Biobank, when PRS is obtained using 10-fold CV, where the P-value threshold for PRS training is fixed at 1; this shows though there is little pc-CE -log10(P values) inflation, the CV-induced artefact is still present. **(G)** Schematic of the Mundlak correction, where the means of chromosomal PRS per fold in a 10-fold CV are obtained and used as covariates in CE tests. **(H)** pc-CE -log10(P values) on LifetimeMDD (black) and CIDI-SF derived symptoms of MDD (green) in UK Biobank, when PRS is obtained using 10-fold CV and pc-CE is performed with Mundlak correction. **(I)** wg-CE -log10(P values) on LifetimeMDD (black) and CIDI-SF derived symptoms of MDD (green) in UK Biobank, when PRS is obtained using 10-fold CV and pc-CE is performed with Mundlak correction.

**Extended Data Figure 4:**
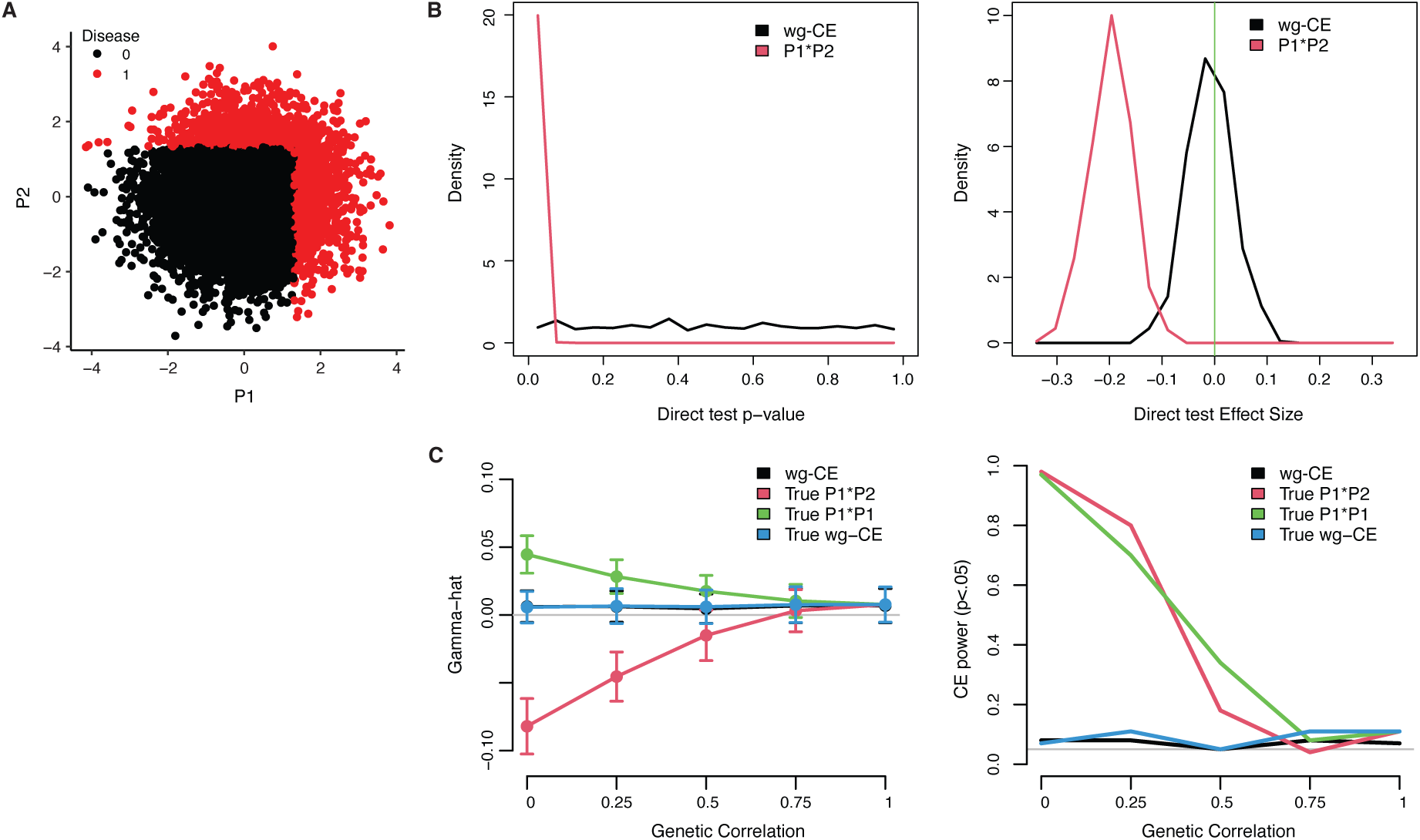
**(A)** Simulated liabilities of subtypes of a disease (P1 and P2) where the two liabilities are antagonistic (negative 𝛾); cases of the disease are coloured in red; controls are coloured in black. **(B) (Left)** Density of P values of wg-CE tests performed on PRS obtained for the disease (black line) and cd-CE tests performed on PRS obtained on the simulated subtypes (P1 * P2, red line). **(Right)** density of 𝛾 estimates obtained from the two tests in the left panel. **(C) (Left)** Estimates of 𝛾 in wg-CE test using simulated effects on disease (black), cd-CE test using simulated P1 and P2 effects (red), wg-CE test for simulated P1 effects (P1 * P1, green), and wg-CE test using estimated effects on disease (blue). Points show averages across 100 simulation replicates, and error bars indicate +/-1 standard deviation. The x-axis varies the genetic correlation between simulated P1 and P2. **(Right)** Power to detect significant 𝛾 in wg-CE and cd-CE tests; the grey line indicates the nominal P value = 0.05 threshold; power to detect significant CE decreases as genetic correlations between P1 and P2 increases.

## Author contributions

JR, JF, AD and NC wrote the paper. NC designed the study. MK performed PA-FGRS analyses. JM supported the Mundlak correction. KGH, TW, AB and AJS supported the iPSYCH analyses. A list of all iPSYCH Consortium members are provided in **Supplementary Table S22**. LH supported the UKB analyses. AD performed the simulations. JR performed all other analyses. NC supervised the study. All authors reviewed the paper.

## Acknowledgements

AJS, AD, JF, KSK and NC are supported by R01MH130581 from the NIH. AJS is supported by Lundbeckfonden Fellowship R335-2019-2318. The iPSYCH team is supported by grants from the Lundbeck Foundation (R102-A9118, R155-2014-1724, and R248-2017-2003), NIMH (1R01MH124851-01) and the Universities and University Hospitals of Aarhus and Copenhagen. The Danish National Biobank resource is supported by the Novo Nordisk Foundation. High-performance computer capacity for handling and statistical analysis of iPSYCH data on the GenomeDK HPC facility is provided by the Center for Genomics and Personalized Medicine and the Centre for Integrative Sequencing, iSEQ, Aarhus University, Denmark. The authors gratefully acknowledge the support of all collaborators and participants in UKB, iPSYCH, and PGC cohorts who made this work possible. The authors also thank Noah Zaitlen, Bertram Müller-Myhsok and Bastian Rieck for valuable discussions.

## Ethical approval

This research was conducted under the ethical approval from the UK Biobank Resource under application no. 28709 and 163937. The use of iPSYCH data follows standards of the Danish Scientific Ethics Committee, the Danish Health Data Authority, the Danish Data Protection Agency, and the Danish Neonatal Screening Biobank Steering Committee. Data access was via secure portals in accordance with Danish data protection guidelines set by the Danish Data Protection Agency, the Danish Health Data Authority, and Statistics Denmark.

## Data availability

UK Biobank genotype and phenotype data used in this study are from the full release (imputation version 2) of the UK Biobank Resource obtained under application no. 28709 and 163937. We used publicly available summary statistics from PGC29 and 23andMe from the Psychiatric Genomics Consortium (https://www.med.unc.edu/pgc/results-and-downloads), with references in **Supplementary Table S5**. Individual-level Danish data is not publicly available due to institutional restrictions on data sharing and privacy concerns. Summary statistics of the GWAS on the gSEM common factor are available at: https://doi.org/10.6084/m9.figshare.28652153.

## Code availability

Publicly available tools that are used in data analyses are described wherever relevant in Methods and Reporting Summary, in particular CE is available at https://github.com/nadavrap/CoordinatedInteractions as previously published; custom code implementing CE in this paper is available at: https://github.com/caina89/psychCE.

