## SupplementaryMaterials for "Genetic risk effects on psychiatric disorders act in sets"

### Supplementary Methods

### UKBiobank

#### Sample filtering in UK Biobank

Of all 502,637 samples in UKBiobank[1] full release, we performed the following QC steps to select the samples for use in our analyses. We first removed samples that were not included in the UKBiobank full release PCA analysis, which includes samples that were indicated as "het.missing.outliers" ("Indicates samples identified as outliers in heterozygosity and missing rates, which indicates poor-quality genotypes for these samples"), "excess.relatives" ("Indicates samples which have more than 10 putative third-degree relatives in the kinship table"), and whose "Submitted.Gender" were different from "Inferred.Gender". Applying these filters brought the sample size down to 407,219. We checked that the remaining sample contains only one out of any pair or group of related individuals with relatedness > 0.05. We then selected samples indicated to be "in.white.British.ancestry.subset" ("Indicates samples who self-reported 'White British' and have very similar genetic ancestry based on a principal components analysis of the genotypes"), resulting in a sample size of 337,545.

We then removed 337 samples indicated as having "putative.sex.chromosome.aneuploidy" ("Indicates samples identified as putatively carrying sex chromosome configurations that are not either XX or XY"). Finally, we removed 79 samples who have withdrawn their consent for use of their genetic data in analyses, arriving at our final set of 337,129 samples passing QC1. Of these samples, 37,041 were part of UK Biobank Lung Exome Variant Evaluation (UKBiLEVE), a study for chronic obstructive pulmonary disease (COPD)[2]. We retain all samples in UKBiLEVE, but as they are genotyped using a custom array optimised for coverage over regions implicated in lung health and disease, we consistently use 'genotyping array' as a covariate in all our analyses.

#### Genotype quality control

We performed stringent filtering on imputed variants (version 3) used for GWAS in this study, removing all insertions and deletions (INDELs) and multi-allelic SNPs: we hard-called genotypes from imputed dosages at 9,720,420 biallelic SNPs with imputation INFO score greater than 0.9, MAF greater than 0.1%, and P value for violation of Hardy-Weinberg equilibrium > $10^{-6}$, in individuals with a genotype probability threshold of 0.9 (individuals with genotype probabilities below 0.9 would be assigned a missing genotype). Of these, 5,776,313 SNPs are common (MAF > 5%). We consistently use these SNPs for all analyses in this study.

We performed principal component analysis (PCA) on directly genotyped SNPs from samples in UKBiobank and used PCs as covariates in all our analyses to control for population structure using flashPCA[3]. From the array genotype data, we first removed all samples who did not pass QC, leaving 337,129 White-British, unrelated samples. We then removed SNPs not included in the phasing and imputation and retained those with minor allele frequencies (MAF) >= 0.1%, and P value for violation of Hardy-Weinberg equilibrium > $10^{-6}$, leaving 593,300 SNPs. We then removed 20,567 SNPs that are in known structural variants (SVs) and the major histocompatibility complex (MHC) as recommended by UKBiobank[1], leaving 572,733 SNPs. Of these, 334,702 are common (MAF > 5%), and from these common SNPs we further filtered based on missingness <0.02 and pairwise LD r2 < 0.1 with SNPs in a sliding window of 1000 SNPs to obtain 68,619 LD-pruned SNPs for computing PCs using flashPCA. We obtained 20 PCs, their eigenvalues, loadings and variance explained, and consistently use these PCs as covariates for all our genetic analyses.

**iPSYCH**

**Cohort description**

We conducted GWAS on two independent cohorts from the Integrative Psychiatric Research Consortium (iPSYCH) cohort (2012 and 2015i).

The Lundbeck Foundation initiative for Integrative Psychiatric Research (iPSYCH)[4,5] is a case-cohort study of all singleton births between 1981 and 2008 to mothers legally residing in Denmark and who were alive and residing in Denmark on their first birthday (N=1,657,449). The iPSYCH 2015 case-cohort comprises two enrollments from this base population. The iPSYCH 2012 case-cohort enrolled 86,189 individuals (30,000 random population controls; 57,377 psychiatric cases)[4]. The iPSYCH 2015i case-cohort expanded enrollment by an additional 56,233 individuals (19,982 random population controls; 36,741 psychiatric cases)[4,5]. DNA was extracted from dried blood spots stored in the Danish Neonatal Screening Biobank[6] and genotyping was performed on the Infinium PsychChip v1.0 array (2012) or the Global Screening Array v2 (2015i). Psychiatric diagnoses were obtained from the Danish Psychiatric Central Research Register (PCR)[7] and the Danish National Patient Register (DNPR)[8]. Diagnoses in these registers are made by licensed psychiatrists during in- or out- patient specialty care but diagnoses or treatments assigned in primary care are not included. Linkage across population registers, to parents where known, and to the neonatal biobank is possible via unique citizen identifiers of the Danish Civil Registration System[9].

The use of this data follows standards of the Danish Scientific Ethics Committee, the Danish Health Data Authority, the Danish Data Protection Agency, and the Danish Neonatal Screening Biobank Steering Committee. Data access was via secure portals in accordance with Danish data protection guidelines set by the Danish Data Protection Agency, the Danish Health Data Authority, and Statistics Denmark.

**Sample selection and genotype quality control**

For this study, we use an unrelated, homogeneous ancestry subset of these data from the iPSYCH2012[4] and iPSYCH2015i cohorts[5].

Genotype phasing, imputation, and quality control were performed in parallel in the iPSYCH2012 and iPSYCH2015i cohorts according to custom, mirrored protocols. Briefly, phasing and imputation were conducted using BEAGLEv5.1[10,11], both steps including reference haplotypes from the Haplotype Reference Consortium v1.1 (HRC)[12]. Quality control was applied prior to and following imputation to correct for missing data across SNPs and individuals, SNPs showing deviations from Hardy-Weinberg equilibrium in controls, abnormal heterozygosity of samples, genotype-phenotype sex discordance, minor allele frequency (MAF), batch artifacts, and imputation quality. Kinship was detected within and across iPSYCH2012 and iPSYCH2015i cohorts using KING[13], censoring to ensure no second degree or higher relatives remained. Ancestry was examined using the smartpca module of EIGENSOFT[14], and PCA outliers from the set of iPSYCH were excluded.

Using 5,210,642 and 5,222,714 SNPs (MAF >= 0.05, Beagle DR2 >= 0.9, P value for HWE violation > $10^{-6}$) on 42,250 individuals in iPSYCH2012 and 23,351 individuals in iPSYCH 2015i respectively, we calculated PRS for each of the 15 summary statistics (phenotyped, imputed and MTAG GWAS) using PRSice v2[15], using the options --clump-kb 250kb --clump-p 1 --clump-r2 0.1 --interval 5e-05 --lower 5e-08. We used the top 25 genomic PCs from all 42,250 and 23,351 individuals in iPSYCH2012 and iPSYCH2015i respectively as covariates to control for population structure in each of the cohorts.

#### Psychiatric disorder phenotypes

iPSYCH was ascertained for five major psychiatric disorders: Major Depressive Disorder (MDD) , Bipolar Disorder (BPD), Schizophrenia (SCZ), Attention Deficit/Hyper-activity Disorder (ADHD) and Autism (AUT). Cases were ascertained using the Danish National Patient Register[16] as well as the Danish Psychiatric Central Research[7]. As a result, individuals were either in- or out-patients at a hospital. As first-line general practitioners in Denmark are able to also treat some psychiatric disorders, iPSYCH is thus biased towards more severe cases. Sample sizes or each of the five disorders included are shown in **Supplementary Table S1**.

For individual disorders in iPSYCH, we define cases as individuals having a diagnosis for the disorder of interest, and controls as all individuals without the disorder, including those might have another disorder.

We define two different definitions of comorbidity in disorder pairs in iPSYCH: the "*Both*" and the "*Any*" phenotypes. Cases in the *Both* phenotype are individuals who have diagnostic codes for both disorders in a disorder pair; cases in the *Any* phenotype are individuals who have diagnostic codes for either or both disorders in the disorder pair. For all comorbidity phenotypes, we use a consistent set of control individuals from the iPSYCH2015 cohort drawn from the general population without any of the mental disorders ANO, SCZ, MDD, AUT, ADHD, BPD. This is fully described in **Figure 4** and **Supplementary Table S1**.

Notably, for all PRS-based analyses on comorbidity phenotypes, we only perform them in the five disorder pairs with more than 250 cases in the *Both* phenotype in both iPSYCH cohorts (ADHD-

MDD, AUT-ADHD, AUT-MDD, BPD-MDD, SCZ-MDD; **Supplementary Table S13**). For all PA-FGRS-based analyses (see section "Pearson-Aitkens Family Genetic Risk Scores") that are performed on the combined iPSYCH2015 dataset, which in inclusive of both iPSYCH cohorts, we perform them on all disorder pairs (**Supplementary Table S13**).

#### Danish register sample description

In Denmark in 1968, all individuals residing and alive in Denmark were registered into the Danish Register[7] for administrative purposes by the Danish Civil Registration System (CRS). This data has been made available to Danish researchers and its link with medical registers can be utilized to extract information on Danish individuals, as was done for iPSYCH.

We use the Danish Register to define new comorbidity phenotypes, using the same definitions as we had for the iPSYCH cohort for both the *Any* and the *Both* phenotypes. Sample sizes specific to these comorbidity phenotypes in the Danish Register can be found in **Supplementary Table S13**.

#### Pearson-Aitkens Family Genetic Risk Scores (PA-FGRS)

#### PA-FGRS estimates of individual disorders

Pearson-Aitkens Family Genetic Risk Scores (PA-FGRS)[17,18] estimates the genetic liability of individuals through use of extended family information, linked through genealogies or family trees. It calculates genetic liability scores from input genealogy data from all available relatives (21 on average). The method[17] builds on a liability threshold model, where all individuals are assumed to have a latent liability that has a genetic and an environmental component, and where the genetic component has a covariance structure given by the kinship matrix. Under this model, the observed disease status reflects if an individual is above or below the threshold. This means that, given that a relative is observed to have the disorder, expected liability of the proband changes away from the population mean. The method updates the expected liability of the proband by giving each affected or unaffected family member, and adjusts the liability score accordingly.

The calculation of individual disorder PA-FGRS takes as input the date of onset or date of end for follow-up for the specific trait or disease of interest and takes into account various measures[17]: Leveraging the population-wide representation of the registers, we estimate sex and birth year specific cumulative incidence proportions. PA-FGRS initiates as a Gaussian distribution of risk with a mean of 0 and standard deviation of 1 for each target individual, and an adjusted liability per individual is derived based on the case status and time to end-of-follow-up for each of the relatives, as well as population prevalence of disorder.

#### PA-FGRS estimates of comorbidity phenotypes

PA-FGRS scores for comorbidity phenotypes are using genealogy data in the Danish Register, using the same definitions in iPSYCH as described above. There are several noteworthy and technical points to consider regarding the calculation of PA-FGRS for comorbidity phenotypes. For the *Both* definition, this is done by treating all relatives with both disorders as affected and all other relatives as unaffected. For the *Any* definition, this is done by treating all relatives with one or both the disorders as affected and all relatives without as unaffected. To adapt this to the liability threshold model, we set the threshold corresponding to the empirical prevalence, treat all unaffected as completely observed and arbitrarily set the heritability parameter to 0.40.

Notably, we essentially make the assumption that comorbidity liability is independent of age for all comorbidity phenotypes we define in this paper, as we do not consider the order in which the two diagnoses in each disorder pair are made.

**PA-FGRS vs PRS**

We observe a big power difference in PRS vs PA-FGRS in our CE tests. We think this may be due partly to the massive genealogy involved in obtaining PA-FGRS as compared to the small reference GWAS sample sizes in GWAS where we obtain PRS from, but may also be indicative of the differences between using family-based liability estimates and additive genetic liability scores.

As PA-FGRS is estimated using disorder status of relatives of individuals, it encompasses more than just the additive genetic effects PRS captures, but also the shared environment[19], indirect genetic effects[20,21] and non-additive genetic effects. Assortative mating, which has been shown to be prevalent among psychiatric disorders[22], will have direct effects on PA-FGRS estimates. However, it has been previously shown that assortative mating (which can be identified using non-zero correlations between the PRS used[23]) does not cause false positive CE in simulations[24]. So even if PA-FGRS on psychiatric phenotypes are, potentially, capturing much more assortative mating than PRS, they would not be biasing CE findings. Likewise, population structure does cause false positive CE, but simulations have shown that this can be mitigated well using PCs as covariates in practice.

As such we think the big differences in power between PRS and PA-FGRS in CE is due to sample size differences in the training data, and PA-FGRS captures more types of liabilities than PRS does.

**Coordinated Epistasis (CE) considerations and interpretations**

**CE models**

The CE framework as proposed by Sheppard et al.[24] uses PRS as proxies for synergistic sets, and tests for significant non-zero interaction effect ($\gamma$) using a log likelihood ratio test. Sheppard et al. first partition the genome into two sets of even- and odd-numbered chromosomes and create PRS for each set (EO-CE, equation 1).

$$y \sim \alpha_o \ PRS_o + \alpha_e \ PRS_e \ + \ \gamma_{eo} PRS_o * PRS_e \qquad [1]$$

Where y = phenotype outcome, $PRS_o$ = odd-partition polygenic risk score, $\alpha_o$ = effect estimate of the additive contribution of $PRS_o$, $\gamma_{eo}$ = interaction effect estimate, indicative of direction of effect of the interaction. Sheppard et al. also demonstrated that partitioning the genome by chromosome and testing for CE across all chromosome pairs in a joint F test improves power (pc-CE, equation 2).

$$y \sim \alpha_i \ PRS_i + \ldots + \alpha_j \ PRS_j \ + \sum_{i \neq j}^{1 \to 22} \gamma_{ij} (PRS_i * PRS_j) \qquad [2]$$

Where $PRS_i$ represents the PRS of an individual chromosome, and $\gamma_{ij}$ captures the interaction effect between each of the chromosome pairs. We report the mean of this value (mean($\gamma_{ij}$)) and the standard error associated with this mean for all pc-CE analyses.

In our work, we extend the CE framework and substitute even and odd-numbered partitioned PRS with whole-genome PRS or the PA-FGRS score (wg-CE and fgrs-CE, equation 3).

$$y \sim \alpha PRS \quad + \gamma_{gs} (PRS)^2 \qquad [3]$$

Here, $\gamma_{gs}$ represents the interaction estimate of using either genetic score (gs) interactions: whole-genome PRS with itself, or PA-FGRS with itself (within-phenotype CE).

For all analyses, even though we test on binary phenotypes, we perform linear, logit and probit regressions to identify interaction effects on different scales.

**CE and previously published models of comorbidity**

Interpretations of CE results are as follows: if $\gamma$ = 0, all genetic effects on the disease are additive. If $\gamma$ < 0, certain genetic effects co-occur less in cases than expected under additivity, and if $\gamma$ > 0 then some genetic effects co-occur more in cases than expected under additivity, constituting "synergistic sets". When genetic effects from different synergistic sets would show antagonism with each other in CE tests.

These models of antagonism and synergism can be aligned with models of comorbidity that are previously described by Klein and Riso[25] and Neale and Kendler[26]. Specifically, they describe six models: (i) there is an underlying liability shared between the two disorders; (ii) having one disorder drastically increases the risk for the second disorder (random multiformity); (iii) having high liability on one disorder increases the risk for the second disorder (extreme multiformity); (iv) the two disorders and the comorbidity can be seen as separate disorders with their own liability; (v) disorder one liability is correlated with disorder two liability; and (vi) one disorder causes the other disorder (direct causal models).

The additive, synergistic and antagonistic architectures CE distinguishes between align with these comorbidity models: the model where there is a single underlying shared liability for comorbidity, distinct from the liabilities of the two constituting disorders (i) is consistent with the additive architecture. The model where comorbidity liability is made of two distinct disorders liabilities (iv) is consistent with the antagonistic architecture. We find this to be the case for comorbidities in two disorder pairs (ADHD-MDD, BPD-MDD), and these findings also support current nosological distinctions between the disorders in the pair. If one disorder drastically increases the risk for the second disorder (ii and iii) would both align with the synergistic architecture, where risk effects of both disorders, together, form a "synergistic set". In this case, whether the two disorders should remain classified as different disorders nosologically should be re-evaluated. We do not observe this between any of the disorder pairs we examine in this paper.

Correlation between liability of the two disorders in question (v) is what we currently estimate as genetic correlation (rG)[27], and is accounted for in CE tests, such that any determination of additive, antagonistic or synergistic architectures are robust across all values of rGs (**Extended Data Figure 4**). In fact, our results show that the level of rG between genetic effects for constituents disorders for a comorbidity is insufficient in determining its architecture. This observation may explain the inconsistency between observed rates of comorbid psychiatric disease within families and genetic correlations estimated from SNP data.

Finally, causation between two disorders (vi) is not explored in this paper due to the requirement of causal inference, which is beyond the scope of this paper.

**Incremental MTAG considerations**

We propose two ways MTAG[28] may identify a CE signal in its focal GWAS (in this case, for LifetimeMDD): it can do so through increasing power to identify CE in the focal GWAS, or capture potential CE between the focal GWAS and other input GWAS. If it was the former, we expect to see increasing power (lower P values) to identify the same CE finding (similar mean $\gamma$ estimates) with increasing effective sample size ($N_{eff}$) if MTAG was performed incrementally using increasing number of input GWAS summary statistics (incrementalMTAG, **Methods, Extended Data Figure 2**) in MTAG.all. Any input GWAS that introduce CE not native to the focal phenotype LifetimeMDD, consistent with the latter explanation, may result in unpredictable changes in P-values and $\gamma$ estimates inconsistent those due to increasing power due to a higher $N_{eff}$.

In performing performing MTAG incrementally (incrementalMTAG) using an increasing number of input GWAS summary statistics inputs for MTAG.all, ordering them based on their absolute genetic correlations (rG) with LifetimeMDD, we find that the results are largely consistent with the former explanation, with the exceptions of shallow definitions of depression as well as the Townsend deprivation index (**Supplementary Table S4, Extended Data Figure 2**). While these result in lower power to detect a negative mean $\gamma$ estimate, or result in a positive mean $\gamma$ estimate, input GWAS on family history depression or measures of stress and neuroticism in incrementalMTAG result in increasingly higher power to identify a negative mean $\gamma$ estimate, consistent with the first hypothesis that they merely increase power in identifying CE effects in LifetimeMDD.

#### Using PRS as an interaction term within-cohort

We obtain PRS in a few different ways (i) using external summary statistics, e.g. from PGC or iPSYCH2012 to obtain PRS in iPSYCH2015i, and (ii) using internal summary statistics, through the use of a 10-fold cross validation (10xCV) approach.

#### Cross-validation (CV) within a cohort

The latter 10xCV approach was performed as follows. We partitioned the input phenotype individuals into ten non-overlapping test sets of 10% of the total sample size of that phenotype. The remaining 90% are training, which were used to obtain discovery GWAS summary statistics using plink. Clumping and Thresholding (C+T) is performed on the 90% GWAS summary statistics per fold, and PRS is obtained for the 10% test sets using the individual-level genotypes and effects at the C+T loci from the 90% GWAS. PRS from all 10 folds are concatenated to give PRS for the whole cohort, to be used within the CE tests (**Extended Data Figure 3**).

#### Artefact using CV PRS in CE analysis

10xCV can introduce artefacts into the CE test through the following:

Different GWAS was used in generating the PRS: the 90% samples GWAS performed for generation of PRS in each 10% test set are different. This difference is enough to generate “batch” effects in PRS obtained between test sets. This difference propagates through the C+T and best P-value threshold selection steps of PRS generation implemented in PRSice, resulting in different SNPs and best P-values thresholds selected for the different folds.

These “batch” effects create artificial correlations between PRS from different chromosomes, which will result in spurious interaction effects in CE tests. This “batch” effect will be present regardless of the number of folds one uses in CV (**Extended Data Figure 3**).

Mitigating steps such as performing C+T on an external GWAS (or a 100% sample GWAS) and setting a constant P-value threshold to be used across all folds are performed can remove spurious CE signal, though the remaining slight differences in effect sizes in each 90% GWAS are still sufficient to generate the “batch” effects. This can be detected through the artificial correlation between PRS between chromosomes due to the CV-induced “batch” effect that persists even when no spurious CE is identified (**Extended Data Figure 3**).

#### Mundlak

The Mundlak method was originally described for use in the field of econometrics[29]. We adapt the method here to correct for a batch effect introduced by Cross-Validation (CV) used in obtaining

PRS within the CE framework. Specifically, we implement the method in line with the description by Rabe-Hesketh and Skrondal[30]. **Extended Data Figure 3** outlines how we perform the Mundlak correction within the CE framework.

First, we obtain the mean value across PRS for each individual fold. Next, we subtract this mean from each per-fold, per-chromosome PRS subset. Last, within the CE mathematical equations, we include a variable that lists for each individual the specific mean that it was adjusted to (see equation below). As such, the Mundlak correction will correct for the batch effect by keeping the variation within batches but eliminating the variation between batches.

$$y \sim \alpha_1 \; PRS'_1 + \alpha_2 \; PRS'_2 \; + \; \mu_f \; + \; \gamma_{1,2} PRS'_1 * PRS'_2 \qquad [4]$$

Where y is the outcome phenotype, $PRS'_1$ is the fold-means adjusted PRS of genomic partition 1 where $PRS'_1 = PRS_1 - \widehat{PRS_1}$, $PRS'_2$ is the fold-means adjusted PRS of genomic partition 2, $\mu_f$ is a vector of fold means and $\gamma_{1,2}$ is the Mundlak corrected resulting CE interaction effect estimate. This adjustment extends to each of the different genomic partitions used to obtain PRS for the CE framework.

We performed the Mundlak correction on LifetimeMDD to showcase the result of correcting for the artifact in this manner is substantial (**Extended Data Figure 3**).

**Supplementary Table Legends**

*Supplementary tables are provided in separate Excel file.*

**Supplementary Table S1: Phenotype definitions, sample size and prevalence of individual disorders in iPSYCH and UK Biobank.** This table describes all phenotypes we use in UKBiobank and iPSYCH2015, including their sample sizes, case prevalence, binary notations, and where relevant, source references for publications with their descriptions and characterizations. For all iPSYCH phenotypes we include the ICD-10 codes used to identify individuals with the respective disorders.

**Supplementary Table S2: Detailed description of phenotypes that are used as input into the incremental MTAG analyses of the MTAG.All phenotype.** This table lists the 12 phenotypes that are used, in addition to the LifetimeMDD phenotype listed in **Supplementary Table S1**, as input GWAS for the incremental MTAG analyses. The table includes phenotype definitions, binary notations, sample sizes and prevalences of these phenotypes and describes on which phenotypes were included in which MTAG meta-analyses. Of note, this table is derived from a previous publication where the MTAG meta-analyses are described and characterized[31].

**Supplementary Table S3: CE statistics for LifetimeMDD and for each previously published MTAG meta-analyses with LifetimeMDD as outcome in the UK Biobank.** This table reports pc-CE and wg-CE test statistics for analyses performed in the UK Biobank using Mundlak correction (**Supplementary Methods**), including CE tests performed with linear regression (lm), logistic regression with logit link function (logit) and logistic regression with probit link function (probit, **Methods**). For pc-CE statistics, mean $\gamma$ estimates across 231 chromosome-pairs (meangamma) and their standard errors (meangammaSE) are reported; for wg-CE single $\gamma$ estimates (gamma) and their standard error (gammaSE) are reported; P values (pvalue) are obtained using a log-likelihood ratio test (LRT) and adjusted using Benjamini-Hochberg (BH) correction (padj); upper (uci) and lower (lci) bounds of the 95% confidence interval are reported for each CE analysis.

**Supplementary Table S4: CE statistics for LifetimeMDD using incremental MTAG summary statistics.** This table reports the resulting statistics from performing CE analyses on LifetimeMDD using PRS on derived from incrementalMTAG summary statistics where LifetimeMDD is the focal phenotype, where increment indicates the number of input GWAS used in incrementalMTAG, and incr.GWAS indicates last GWAS added to incrementalMTAG. We show pc-CE and wg-CE test statisticsfor CE tests performed with linear regression (lm), logistic regression with logit link function (logit) and logistic regression with probit link function (probit). For pc-CE statistics, mean $\gamma$ estimates across 231 chromosome-pairs (meangamma) and their standard errors (meangammaSE) are reported; for wg-CE single $\gamma$ estimates (gamma) and their standard error (gammaSE) are reported; P values (pvalue) are obtained using a log-likelihood ratio test (LRT); upper (uci) and lower (lci) bounds of the 95% confidence interval are reported for each CE analysis.

**Supplementary Table S5: Reference of PGC summary statistics used when obtaining external PRS in iPSYCH.** This table lists the references and sample size information of the five psychiatric disorder summary statistics from the Psychiatric Genetics Consortium (PGC) and the specific publications they were obtained from. Importantly, the summary statistics listed here did not include iPSYCH participants to avoid overfitting in training PRS in the iPSYCH cohorts. Disorders included are: Attention-Deficit/Hyperactivity Disorder (ADHD), Autism (AUT), Bipolar Disorder (BPD), Major Depressive Disorder (MDD) and Schizophrenia (SCZ).

**Supplementary Table S6: CE test statistics for five psychiatric disorders in iPSYCH using summary statistics from the PGC and MTAG performed on PSYCH and PGC summary statistics.** For each of the five disorders (Attention-Deficit/Hyperactivity Disorder (ADHD), Autism (AUT), Bipolar Disorder (BPD), Major Depressive Disorder (MDD) and Schizophrenia (SCZ)) discussed in this paper, this table lists pc-CE (per-chromosome) and wg-CE (whole-genome) test statistics. Input summary statistics listed in PRS are either from iPSYCH2012, iPSYCH2015i, PGC or meta-analyzed using MTAG using both iPSYCH sub-cohorts and alternating the focal phenotype between iPSYCH and PGC. We show pc-CE and wg-CE test statistics for CE tests performed with linear regression (lm), logistic regression with logit link function (logit) and logistic regression with probit link function (probit). For pc-CE statistics, mean $\gamma$ estimates across 231 chromosome-pairs (meangamma) and their standard errors (meangammaSE) are reported; for wg-CE single $\gamma$ estimates (gamma) and their standard error (gammaSE) are reported; P values (pvalue) are obtained using a log-likelihood ratio test (LRT) and adjusted using Benjamini-Hochberg (BH) correction (padj); upper (uci) and lower (lci) bounds of the 95% confidence interval are reported for each CE analysis.

**Supplementary Table S7: CE test statistics of cross-cohort CE tests between iPSYCH sub-cohorts and PGC.** This table reports CE test statistics for pc-CE (per-chromosome) and wg-CE (whole-genome) tests performed to test for CE between iPSYCH sub-cohorts iPSYCH2012 or iPSYCH2015i and the PGC summary statistics (**Supplementary Table S5**). We show the input PRS (PRS_A and PRS_B), the pc-CE and wg-CE test statistics for CE tests performed with linear regression (lm), logistic regression with logit link function (logit) and logistic regression with probit link function (probit). For pc-CE statistics, mean $\gamma$ estimates across 231 chromosome-pairs (meangamma) and their standard errors (meangammaSE) are reported; for wg-CE single $\gamma$ estimates (gamma) and their standard error (gammaSE) are reported; P values (pvalue) are obtained using a log-likelihood ratio test (LRT) and adjusted using Benjamini-Hochberg (BH) correction (padj); upper (uci) and lower (lci) bounds of the 95% confidence interval are reported for each CE analysis.

**Supplementary Table S8: Sample size information from the Danish Register for five psychiatric disorders.** This table lists phenotype definitions and sample sizes for five psychiatric disorders in the Danish Register: Attention-Deficit/Hyperactivity Disorder (ADHD), Autism (AUT), Bipolar Disorder (BPD), Major Depressive Disorder (MDD) and Schizophrenia (SCZ). Additionally, register-based and previously published population prevalences[32] are listed. We use the published population prevalences for gSEM analyses.

**Supplementary Table S9: CE test statistics for five disorders using PA-FGRS from the Danish Register.** This table lists the within-phenotype fgrs-CE test statistics on five disorders in iPSYCH, performed using PA-FGRS derived from the genealogy data from the Danish Register. $\gamma$ estimates (gamma) and their standard error (gammaSE) for fgrs-CE tests are reported for CE tests performed with linear regression (lm), logistic regression with logit link function (logit) and logistic regression with probit link function (probit); P values (pvalue) are obtained using a log-likelihood ratio test (LRT) and adjusted using Benjamini-Hochberg (BH) correction (padj); upper (uci) and lower (lci) bounds of the 95% confidence interval are reported for each CE analysis. P values (pvalue) are obtained using a log-likelihood ratio test (LRT) and adjusted using Benjamini-Hochberg (BH) correction (padj).

**Supplementary Table S10: Per-disorder CE test statistics of PRS by FGRS interaction tests.** This table lists the within-phenotype PRS by PA-FGRS CE test statistics on five disorders in iPSYCH. Cohort on which PRS is trained is shown in the PRS column; cohort on which PRS and PA-FGRS is obtained is shown in the Target column; $\gamma$ estimates (gamma) and their standard error (gammaSE) for PRS by PA-FGRS CE tests are reported for CE tests performed with linear regression (lm), logistic regression with logit link function (logit) and logistic regression with probit link function (probit); P values (pvalue) are obtained using a log-likelihood ratio test (LRT) and adjusted using Benjamini-Hochberg (BH) correction (padj); upper (uci) and lower (lci) bounds of the 95% confidence interval are reported for each CE analysis.

**Supplementary Table S11: Cross-disorder wg-CE test statistics for ten disorder pairs in iPSYCH.** This table lists CE test statistics results for tests between PRS of two disorders (disorderA, disorderB) on a focal disorder (Disorder). CE tests are performed on one iPSYCH (Target) cohort using PRS trained using the other iPSYCH cohort or a MTAG result (PRS). We report $\gamma$ estimates (gamma) and their standard error (gammaSE) for PRS by PA-FGRS CE tests are reported for CE tests performed with linear regression (lm), logistic regression with logit link function (logit) and logistic regression with probit link function (probit); P values (pvalue) are obtained using a log-likelihood ratio test (LRT) and adjusted using Benjamini-Hochberg (BH) correction (padj); upper (uci) and lower (lci) bounds of the 95% confidence interval are reported for each CE analysis.

**Supplementary Table S12: Cross-disorder fgrs-CE test statistics for ten disorder pairs in iPSYCH.** This table lists cross-disorder fgrs-CE test statistics for ten disorder pairs in iPSYCH, where the PA-FGRS for the two disorders are shown in the columns FGRS_A and FGRS_B, and the focal disorder is shown in the column Disorder. We report $\gamma$ estimates (gamma) and their standard error (gammaSE) for cross-disorder fgrs-CE tests are reported for CE tests performed with linear regression (lm), logistic regression with logit link function (logit) and logistic regression with probit link function (probit); P values (pvalue) are obtained using a log-likelihood ratio test (LRT) and adjusted using Benjamini-Hochberg (BH) correction (padj); upper (uci) and lower (lci) bounds of the 95% confidence interval are reported for each CE analysis.

**Supplementary Table S13: Comorbidity phenotype definitions and sample size information on the Danish Register in comparison to iPSYCH and its sub-cohorts.** This table lists phenotype definitions, sample sizes and prevalences for the phenotypes *Any* and *Both* defined for all disorder pairs in iPSYCH. We do not take forward comorbidity phenotypes with fewer than 250 cases in individual iPSYCH cohorts for CE analysis.

**Supplementary Table S14: Cross-disorder CE test on comorbidity phenotypes using PRS.** This table reports the test statistics resulting from performing cd-CE tests using PRS of constituent disorders (PRS_A, PRS_B) in all comorbidity phenotypes with more than 250 cases in one iPSYCH cohort (Cohort). CE test is performed in one iPSYCH cohort (cohort) and PRS trained on the other iPSYCH cohort. We report $\gamma$ estimates (gamma) and their standard error (gammaSE) for cross-disorder fgrs-CE tests are reported for CE tests performed with linear regression (lm), logistic regression with logit link function (logit) and logistic regression with probit link function (probit); P values (pvalue) are obtained using a log-likelihood ratio test (LRT) and adjusted using Benjamini-Hochberg (BH) correction (padj); upper (uci) and lower (lci) bounds of the 95% confidence interval are reported for each CE analysis.

**Supplementary Table S15: Cross-disorder CE test on comorbidity phenotypes using PA-FGRS.** This table reports the test statistics resulting from performing cd-CE tests using PA-FGRS of constituent disorders (FGRS_A, FGRS_B) in all comorbidity phenotypes with more than 250 cases in one iPSYCH cohort (Cohort). CE test is performed on the whole iPSYCH data using PA-FGRS obtained from genealogies in the Danish register. We report $\gamma$ estimates (gamma) and their standard error (gammaSE) for cross-disorder fgrs-CE tests are reported for CE tests performed with linear regression (lm), logistic regression with logit link function (logit) and logistic regression with probit link function (probit); P values (pvalue) are obtained using a log-likelihood ratio test (LRT) and adjusted using Benjamini-Hochberg (BH) correction (padj); upper (uci) and lower (lci) bounds of the 95% confidence interval are reported for each CE analysis.

**Supplementary Table S16: Within-phenotype CE statistics for comorbidity phenotypes using PRS.** This table shows the cross-disorder CE test statistics for all comorbidity phenotypes with more than 250 cases in one iPSYCH cohort (cohort), performed using PRS obtained from the other iPSYCH cohort. We report $\gamma$ estimates (gamma) and their standard error (gammaSE) for cross-disorder fgrs-CE tests are reported for CE tests performed with linear regression (lm), logistic regression with logit link function (logit) and logistic regression with probit link function (probit); P values (pvalue) are obtained using a log-likelihood ratio test (LRT) and adjusted using Benjamini-Hochberg (BH) correction (padj); upper (uci) and lower (lci) bounds of the 95% confidence interval are reported for each CE analysis.

**Supplementary Table S17: Within-phenotype CE tests on comorbidity phenotypes using PA-FGRS.** This table shows the cross-disorder CE test statistics for all comorbidity phenotypes with more than 250 cases in one iPSYCH cohort (cohort), performed using PA-FGRS obtained on that iPSYCH cohort. We report $\gamma$ estimates (gamma) and their standard error (gammaSE) for cross-disorder fgrs-CE tests are reported for CE tests performed with linear regression (lm), logistic regression with logit link function (logit) and logistic regression with probit link function (probit); P values (pvalue) are obtained using a log-likelihood ratio test (LRT) and adjusted using

Benjamini-Hochberg (BH) correction (padj); upper (uci) and lower (lci) bounds of the 95% confidence interval are reported for each CE analysis.

**Supplementary Table S18: CE between PRS and PA-FGRS on comorbidity phenotypes.** This table reports the test statistics resulting from performing CE tests between PRS and PA-FGRS for all comorbidity phenotypes with more than 250 cases in one iPSYCH cohort (Cohort), performed using PA-FGRS obtained on that iPSYCH cohort and PRS trained on the other iPSYCH cohort. We report $\gamma$ estimates (gamma) and their standard error (gammaSE) for cross-disorder fgrs-CE tests are reported for CE tests performed with linear regression (lm), logistic regression with logit link function (logit) and logistic regression with probit link function (probit); P values (pvalue) are obtained using a log-likelihood ratio test (LRT) and adjusted using Benjamini-Hochberg (BH) correction (padj); upper (uci) and lower (lci) bounds of the 95% confidence interval are reported for each CE analysis.

**Supplementary Table S19: Within-phenotype CE on individual disorders using PRS derived from gSEM P factor.** This table reports the within-phenotype wg-CE test statistics from CE tests on individual disorders in iPSYCH cohorts (cohort) using PRS obtained from a P factor obtained from a gSEM common factor analysis run on PGC GWAS for five disorders, adjusted for published Danish population prevalences as shown in **Supplementary Table S9**. We report $\gamma$ estimates (gamma) and their standard error (gammaSE) for CE tests performed with linear regression (lm), logistic regression with logit link function (logit) and logistic regression with probit link function (probit); P values (pvalue) are obtained using a log-likelihood ratio test (LRT) and adjusted using Benjamini-Hochberg (BH) correction (padj); upper (uci) and lower (lci) bounds of the 95% confidence interval are reported for each CE analysis.

**Supplementary Table S20: Within-phenotype CE on individual disorders using PRS derived from gSEM P factor and disorder-specific PRS.** This table reports the within-phenotype wg-CE test statistics from CE tests on individual disorders in each iPSYCH cohorts (cohort) using disorder-specific PRS obtained from GWAS on the other iPSYCH cohort and PRS of a P factor obtained from a gSEM common factor analysis run on PGC GWAS for five disorders, adjusted for published Danish population prevalences as shown in **Supplementary Table S9**. We report $\gamma$ estimates (gamma) and their standard error (gammaSE) for CE tests performed with linear regression (lm), logistic regression with logit link function (logit) and logistic regression with probit link function (probit); P values (pvalue) are obtained using a log-likelihood ratio test (LRT) and adjusted using Benjamini-Hochberg (BH) correction (padj); upper (uci) and lower (lci) bounds of the 95% confidence interval are reported for each CE analysis.

**Supplementary Table S21: Within-phenotype CE test on comorbidity phenotypes using PRS derived from gSEM "P" factor.** This table reports the within-phenotype wg-CE and pc-CE test statistics from CE tests on comorbidity phenotypes with case numbers greater than 250 in iPSYCH cohorts (cohort) using PRS obtained from a "P" factor obtained from a gSEM common factor analysis run on PGC GWAS for five disorders, adjusted for published Danish population prevalences as shown in **Supplementary Table S9**. We report $\gamma$ estimates (gamma) and their standard error (gammaSE) for CE tests performed with linear regression (lm), logistic regression with logit link function (logit) and logistic regression with probit link function (probit); P values (pvalue) are obtained using a log-likelihood ratio test (LRT) and adjusted using Benjamini-Hochberg (BH) correction (padj); upper (uci) and lower (lci) bounds of the 95% confidence interval are reported for each CE analysis.

**Supplementary Table S22: Co-authors under the iPSYCH banner and their affiliations.**
